# Urethra contours on MRI: multidisciplinary consensus educational atlas and reference standard for artificial intelligence benchmarking

**DOI:** 10.1101/2025.07.01.25330656

**Authors:** Yuze Song, Lily Nguyen, Anna Dornisch, Madison Baxter, Tristan Barrett, Anders M Dale, Robert T Dess, Mukesh Harisinghani, Sophia C Kamran, Michael A Liss, Daniel JA Margolis, Eric P Weinberg, Sean A Woolen, Tyler M Seibert

## Abstract

**Introduction:** The urethra is a recommended avoidance structure for prostate cancer treatment. However, even subspecialist physicians often struggle to accurately identify the urethra on available imaging. Automated segmentation tools show promise, but a lack of reliable ground truth or appropriate evaluation standards has hindered validation and clinical adoption. This study aims to establish a reference-standard dataset with expert consensus contours, define clinically meaningful evaluation metrics, and assess the performance and generalizability of a deep-learning-based segmentation model.

**Materials and Methods:** A multidisciplinary panel of four experienced subspecialists in prostate MRI generated consensus contours of the male urethra for 71 patients across six imaging centers. Four of those cases were previously used in an international study (PURE-MRI), wherein 62 physicians attempted to contour the prostate and urethra on the patient images. Separately, we developed a deep-learning AI model for urethra segmentation using another 151 cases from one center and evaluated it against the consensus reference standard and compared to human performance using Dice Score, percent urethra Coverage, and Maximum 2D (axial, in-plane) Hausdorff Distance (HD) from the reference standard.

**Results:** In the PURE-MRI dataset, the AI model outperformed most physicians, achieving a median Dice of 0.41 (vs. 0.33 for physicians), Coverage of 81% (vs. 36%), and Max 2D HD of 1.8 mm (vs. 1.6 mm). In the larger dataset, performance remained consistent, with a Dice of 0.40, Coverage of 89%, and Max 2D HD of 2.0 mm, indicating strong generalizability across a broader patient population and more varied imaging conditions.

**Conclusion:** We established a multidisciplinary consensus benchmark for segmentation of the urethra. The deep-learning model performs comparably to specialist physicians and demonstrates consistent results across multiple institutions. It shows promise as a clinical decision-support tool for accurate and reliable urethra segmentation in prostate cancer radiotherapy planning and studies of dose-toxicity associations.

## Introduction

Magnetic Resonance Imaging (MRI) has become an essential tool in the early detection, diagnosis, and treatment of prostate cancer, playing a crucial role in guiding targeted biopsies and facilitating treatment planning^1–7^. Among the anatomical structures visualized on MRI, accurate contouring of the urethra has attracted increasing attention due to its relationship with treatment-related toxicity^8–10^. However, accurate contouring of the urethra on MRI remains a significant clinical challenge for human experts, due to the urethra’s small caliber, anatomical variability, and the limited visibility of its upper half on MRI^11^. Moreover, there is currently no universally accepted guideline or consensus among clinicians regarding the optimal approach to urethral contouring.

Several automated methods based on deep learning and radiomics have been proposed for urethra segmentation, demonstrating promising potential^12–15^. Their reported performance remains difficult to evaluate because there is no accepted contouring guideline, and there are large variations in human physician contours that would serve as the “ground truth” for these models^11^. Moreover, the commonly used Dice score may not be an ideal metric for assessing urethra segmentation. While a Dice Score of 1.0 indicates perfect spatial overlap, the metric heavily penalizes small deviations and over-segmentation, which is problematic given the urethra’s small size and low contrast on MRI. These characteristics can lead to disproportionately low Dice scores even for clinically acceptable segmentations. For example, expert consensus supports sparing the urethra for radiation therapy (RT^9^), but this could be achieved with a urethra avoidance structure that encompasses most of the urethra, even if the boundaries of the contours do not have the sub-millimeter precision that could be required to have a very high Dice Score^16^.

To address these challenges, we constructed multidisciplinary consensus urethra contours for a multi-center dataset, following a similar process to that used in prior work on prostate contouring^11,17^. By incorporating interactive feedback and careful review from multiple experienced clinicians with diverse expertise, we aim to establish a more reliable and confident reference standard. We also proposed an evaluation strategy that combines coverage of the urethra and distance-based deviation metrics, in addition to the traditional Dice Score, which may better reflect clinical relevance and performance in real-world scenarios.

Recognizing the difficulty of the urethra contouring task for many physicians^11^, we also developed a deep learning–based model for urethra segmentation, which we evaluated using the newly constructed expert consensus dataset and the proposed evaluation metrics. We conducted two key comparisons to assess model performance. The first involves a smaller dataset previously used to evaluate human expert annotations^11^, allowing for a direct comparison between the AI model and clinical experts. The second evaluation utilized a larger and more diverse dataset to examine the model’s robustness and generalizability across varied imaging conditions and patient populations. Through this dual evaluation strategy, our goal is to provide a more comprehensive and clinically meaningful assessment of AI performance and to contribute to the development of reliable, AI-assisted tools for prostate cancer RT planning and future studies of RT-related toxicity.

## Methods

### Cases

Two datasets (Dataset 1 and Dataset 2) were utilized for this study. All patients included in these datasets underwent prostate multi-parametric MRI (mpMRI) for the evaluation of known or suspected prostate cancer. Dataset 1 includes four patient cases from Imaging Center 1 and was previously used in a study assessing performance of human experts for prostate contouring and urethra contouring (as well as an AI model for prostate contouring)^11^. Dataset 2 was described in a prior study evaluating automated prostate segmentation tools^17^. This dataset comprises 71 patient cases with MRI acquired on one of eleven scanners at six imaging centers and includes the four cases from Dataset 1 (Table 1). As described previously, cases with substantial imaging artifacts (hip implants, excessive bowel gas, or large body habitus interfering with image quality) were excluded^17^.

**Table 1.** Characteristics of the patient cases included in this study. All n=71 cases are included in Dataset 2. Dataset 1 is a subset of 4 cases from Imaging Center 1, with images acquired on two scanners (GE Healthcare Signa Premier).

| Center and Scanner Platforms for Patient Cases for Validation (n = 71) |  |  |
| --- | --- | --- |
| Cohorts |  |  |
| Imaging Center 1 |  | 13 |
| Imaging Center 2 |  | 14 |
| Imaging Center 3 |  | 12 |
| Imaging Center 4 |  | 10 |
| Imaging Center 5 |  | 10 |
| Imaging Center 6 |  | 12 |
| <u>Institution</u> | <u>MRI scanner models</u> | <u>Number of scanners</u> |
| Imaging Center 1 | GE Healthcare Discovery MRI750,<br>GE Healthcare Signa Premier | 3 |
| Imaging Center 2 | SIEMENS Skyra | 2 |
| Imaging Center 3 | GE Healthcare Signa Premier | 1 |
| Imaging Center 4 | GE Healthcare Signa Premier | 2 |
| Imaging Center 5 | GE Healthcare Discovery MRI750 | 1 |
| Imaging Center 6 | SIEMENS Skyra | 2 |
| Total | 3 models | 11 scanners |

### Reference standard contour development

For both datasets, a panel of four experts was convened to develop the reference standard urethra segmentation dataset^11,17^: two genitourinary (GU) radiologists (with 12 and 21 years’ experience, each surpassing the number of MRIs reported to qualify as prostate MRI experts^18^) and two GU radiation oncologists specializing in MRI-guided prostate radiotherapy (10 years’ experience each). One of the GU radiation oncologists generated initial contours on high-resolution axial *T_2_*-weighted slices^9^. During the panel meeting, all three planes were always visualized, and high-resolution coronal and sagittal *T_2_*-weighted acquisitions were available. Each slice case was reviewed at least twice. All members of the panel in attendance agreed on the final contour for each use.

### Urethra contouring ground rules

The ground rules for the contouring of the urethra were agreed by all the members in the panel^11^. Both the prostatic and membranous urethra were included in the delineation. In cases where the urethra was not clearly visible on a given slice—most commonly at the mid-gland and portions of the prostatic base—the panel reviewed all available anatomic information across the three imaging planes to estimate the most likely location. The urethra was consistently contoured as a continuous structure extending from the bladder neck through the prostatic apex and membranous urethra, terminating at the most superior axial slice where the penile bulb was fully visualized.

### Physician participant eligibility in PURE-MRI study

Eligible participants included radiation oncologists, radiologists, and urologists involved in the diagnosis and/or treatment of prostate cancer. Clinical residents or fellows in these specialties were also eligible if they had completed at least one prostate-focused clinical rotation. All study materials, recruitment communications, and procedures were approved by the Institutional Review Board (IRB) at UC San Diego. Participants were recruited through social media, email outreach, and word-of-mouth within the genitourinary oncology community^11^.

### Auto-segmentation (AI) model development

Training data for the deep-learning AI model included another 151 cases described in a previous publication^19^. One of the radiation oncologist panelists had previously contoured the urethra on these 151 patient cases in 2023 to create sample data for understanding anatomic patterns. These cases were obtained through routine clinical care at Imaging Center 1 on a single 3T scanner (GE Healthcare Discovery 750). Because the exact boundaries of the urethra are often not precisely visible—even when the location of the urethra is discernible—we applied with a 2 mm dilation to the manual contours with OpenCV^20^. The idea was to train the AI model to identify the urethra’s position and course through the prostate rather than necessarily reproduce the exact contours as drawn by the human. The training process utilized nnU-Net^21^, a robust and well-established model architecture. There was a total of 1,000 training epochs. The initial learning rate was 0.1 and gradually decreased to 0.00002 by the final epoch. Training was conducted on a single NVIDIA Tesla V100 GPU at San Diego Supercomputer Center^22^, with a batch size of 8. The input *T_2_*-weighted volumes were cropped to a patch size of 16×320×320 and resampled to a uniform voxel size of 3.0×0.5×0.5 mm³. The upper half of the urethra is almost invisible, which can result in missing segments and produce disconnected components in the segmentation. To address this limitation, a linear interpolation approach was implemented to ensure a continuous segmentation consistent with known anatomical prior information and thus improve overall segmentation performance.

### Evaluation of AI and human performance

We compared urethra segmentations by the AI model to those produced by physician participants in the PURE-MRI study, using the expert consensus urethra contour in Dataset 1 as the reference standard. To evaluate the generalizability and robustness of the AI model, we also assessed its performance on Dataset 2, again using the expert consensus contours as the reference standard. Segmentation performance was quantified using the percent coverage and Dice score for the entire urethra. Percent coverage refers to the percentage of the consensus reference urethra that was included in the AI tool’s (or physican participants’) urethra segmentation. The Dice score ranges from 0 (no overlap) to 1 (perfect overlap), accounting for both perfect and no overlap and penalizes over-segmentation as much as under-segmentation. Percent coverage is a conservative metric in the context of a dose constraint like the near-maximum dose to the urethra. On the other hand, an enormously over-segmentation of the prostate (e.g., the whole prostate) would ensure high percent coverage but would also make it difficult to focally boost dose to prostate cancer. To address this limitation, we also incorporated the maximum 2D Hausdorff distance (Max 2D HD), which captures the maximum deviation between the predicted and reference contours across all axial *T_2_*-weighted slices, thereby providing an indication of degree of over-segmentation.

## Results

### Cases

Patient cases (n=71 in all) used in this study have been described previously^11^. Briefly, Dataset 1 consists of 4 patient cases from a single imaging center, with MRI data acquired using two 3T scanners from one vendor (GE Healthcare)^11^. Dataset 2 consists of 71 cases from six imaging centers, with MRI data acquired using with eleven 3T scanners from two vendors (GE Healthcare; Siemens Healthineers)^17^. None of the cases used an endorectal coil. The panel reached consensus for the urethra segmentation on each slice of all 71 cases. We present all slices for one patient to illustrate a complete urethra volume and serve as a contouring guide/atlas (Supplementary Figure 1). We pick several slices from Supplementary Figure 1 and gave a more detailed annotations about urethra contouring in Supplementary Figure 2. For illustration, we also present the mid axial, coronal and sagittal view for two patients with and without a urinary catheter (Supplementary Figure 3).

### Physician participant contours in PURE-MRI study

As described previously^11^, a total of 62 physicians from 11 countries participated, including oncology, radiology, and urology, with a range of sub-specialization and varying years of experience (Supplementary Table 1). In all, physician participants generated 110 urethra contours.

### Evaluation of AI and human performance on Dataset 1

The AI model generated contours that covered a median 81% of the reference urethra [IQR: 80, 84] for the four cases in Dataset 1 (Table 2). Physician participants’ contouring attempts, in comparison, covered a median 36% of the reference urethra [IQR: 25, 59]. The best-performing group of human participants were those with 5-10 years of experience, who achieved median 53% coverage [IQR: 31, 71].

**Table 2.**
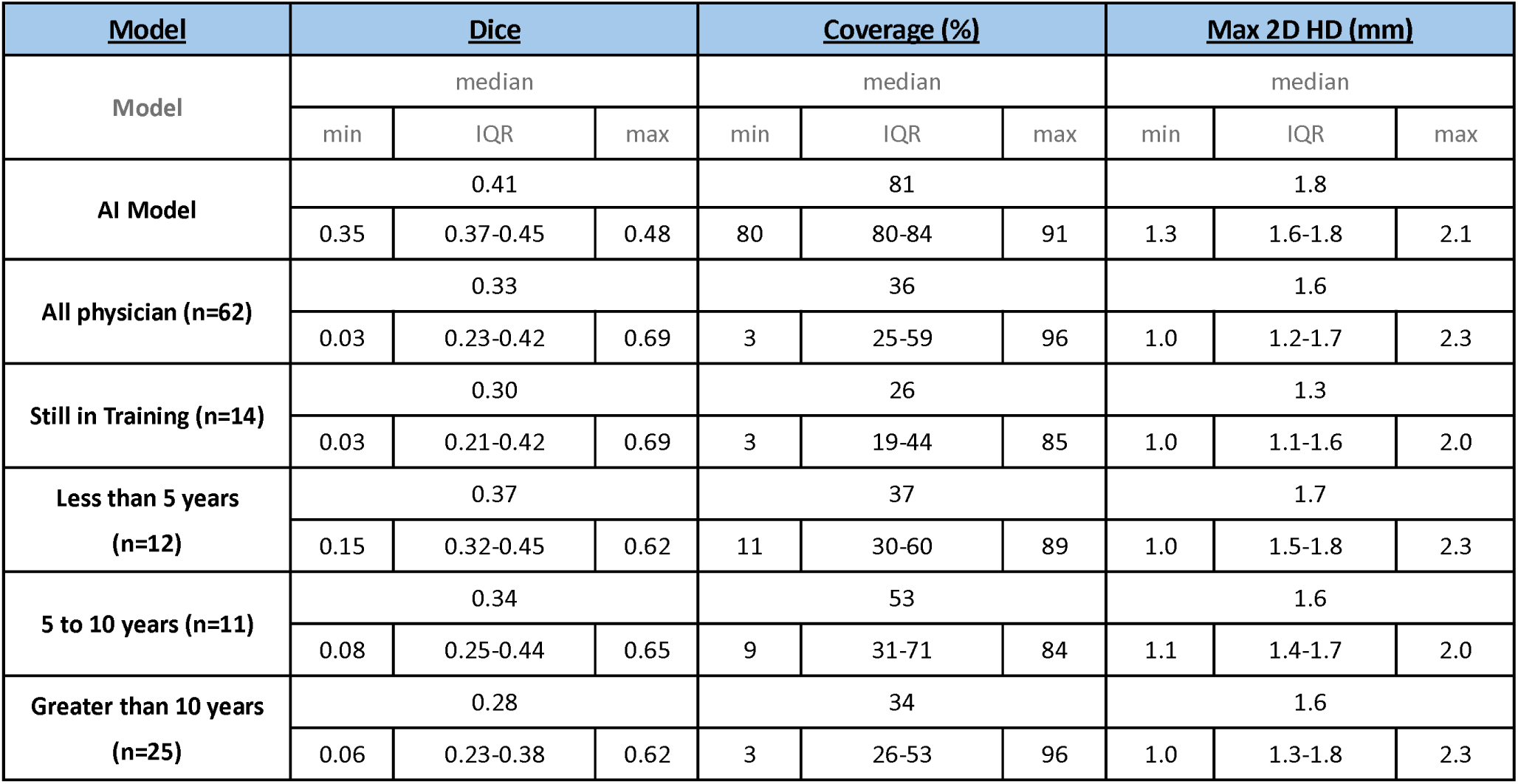
Dice, Coverage (%) and Max 2D HD (mm) of the AI model and physician participants with different year of experience on Dataset 1.

In terms of Dice Score, the AI model achieved a median of 0.41 [0.37, 0.45] across the four cases in Dataset 1 (Table 2). Physician participants had a median Dice of 0.33 [0.23, 0.42]. The subgroup of physicians with <5 years of experience performed best by this metric with median Dice of 0.37 [0.32, 0.45].

The AI model achieved a median Max 2D Hausdorff Distance (HD) of 1.8 mm [1.6, 1.8], meaning a typical AI urethra contour never included prostate tissue more than 1.8 mm from the reference standard (Table 2). Physician participants performed similarly on this metric with median Max 2D HD of 1.6 mm [1.2, 1.7]. The best-performing subgroup was those still in training, who achieved a median of 1.3 mm [1.1, 1.6]. In terms of overall performance range, the AI model produced values for Max 2D HD between 1.3 mm and 2.1 mm, while the range for physician participants extended from 1.0 mm to 2.3 mm.

We present representative *T_2_*-weighted mid axial, sagittal and coronal slices illustrating contours generated by both the AI model and physician participants (Figure 1).

**Figure 1.**
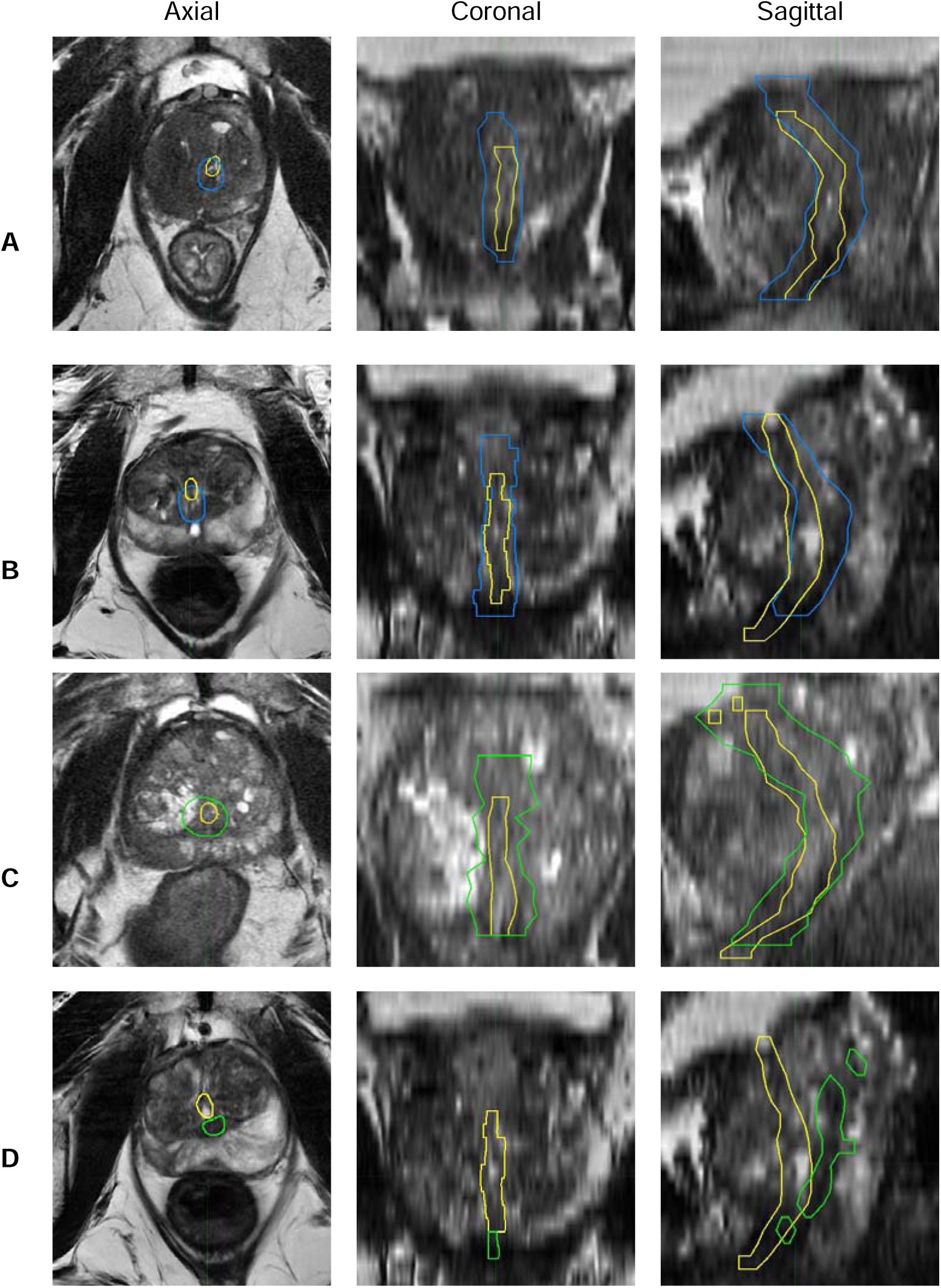
Examples of AI and physician urethra contours, as well as the reference standard (consensus contour by a multidisciplinary panel of four subspecialists. Contours are overlayed on axial, coronal, and sagittal *T_2_*-weighted MRI. The expert-defined urethra contour is shown as yellow contour. The AI generated urethra contour is shown as blue contour. The physician participants’ urethra contours are shown in green. **1A** is an example of AI generated urethra contour with Dice of 0.35, Coverage of Urethra of 91%, and Max 2D HD of 2.1 mm. **1B** is an example of AI generated urethra contour with Dice of 0.38, Coverage of Urethra of 80%, and Max 2D HD of 1.3 mm. **1C** is an example of physician participants’ urethra contour with Dice of 0.21, Coverage of Urethra of 96%, and Max 2D HD of 1.7 mm. **1D** is an example of a physician participant’s urethra contour with Dice of 0.09, Coverage of Urethra of 8%, and Max 2D HD of 1.1 mm.

### Evaluation of AI in Dataset 2

To evaluate the performance of the AI model in a broader range of patients, scanners, and imaging centers, we applied the automated tool to all 71 cases in Dataset 2 (Table 3). Percent coverage of the reference standard urethra was median 89% [IQR: 78, 95]. The broader dataset did reveal some outliers, though, as the worst example was a case with only 34% coverage (Figure 2), and the best was 100% coverage. Median Dice was 0.40 [0.35, 0.44]. Median Max 2D Hausdorff Distance was 2.0 mm [1.5, 2.3], indicating that the AI model’s contour for a typical case strayed no more than 2 mm from the reference standard. In the worst case, the AI model’s contour strayed 2.9 mm from the reference standard in one slice.

**Figure 2.**
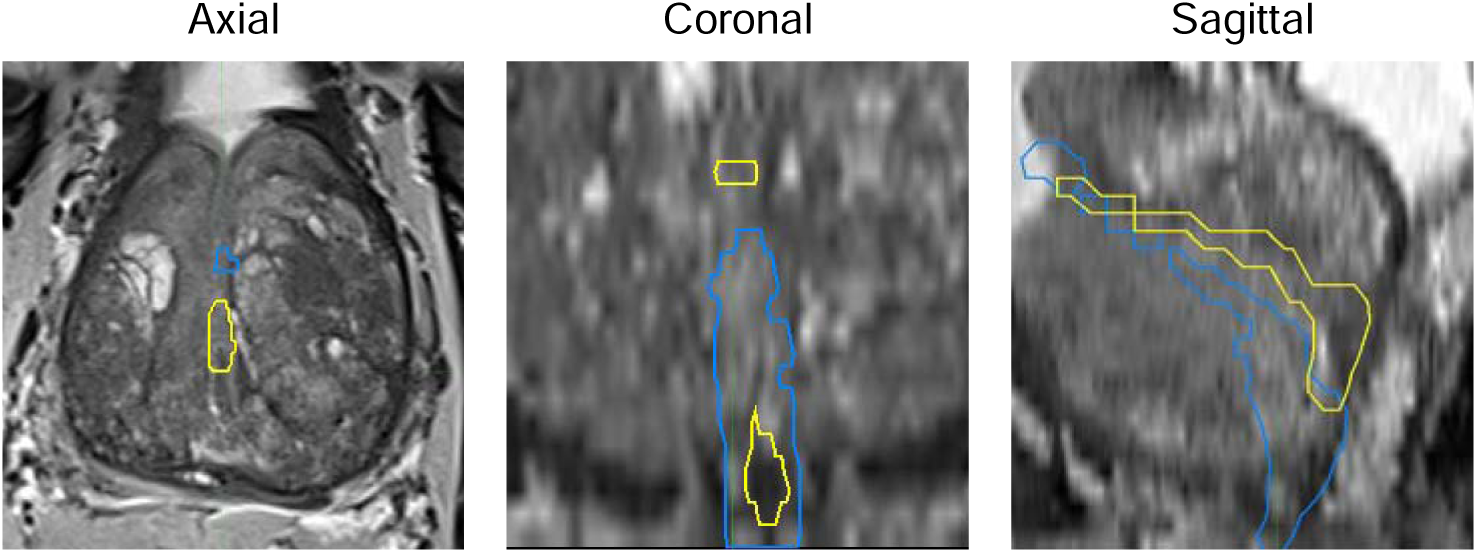
Example auto-segmentation errors of urethra from AI. Contours are overlayed on axial, coronal and sagittal *T_2_*-weighted MRI. The expert-defined urethra contour is shown as yellow contour. The Ai generated urethra contour is shown as blue contour. The AI generated urethra contour with Dice of 0.20, Coverage of Urethra of 34%, and Max 2D HD of 2.9 mm.

**Table 3.** Dice, Coverage (%) and Max 2D HD (mm) of the AI model on Dataset 2.

| <u>Model</u> | <u>Dice</u> |  |  | <u>Coverage (%)</u> |  |  | <u>Max 2D HD (mm)</u> |  |  |
| --- | --- | --- | --- | --- | --- | --- | --- | --- | --- |
| Model | median |  |  | median |  |  | median |  |  |
|  | min | IQR | max | min | IQR | max | min | IQR | max |
| AI Model | 0.40 |  |  | 89 |  |  | 2.0 |  |  |
|  | 0.20 | 0.35-0.44 | 0.54 | 34 | 78-95 | 100 | 1.0 | 1.5-2.3 | 2.9 |

## Discussion

The urethra is a critical organ of interest in prostate cancer treatment because of its relationship to functional outcomes for patients undergoing radiation therapy^8–10^. Measurement of urethra length may also be relevant for predicting outcomes after surgery^23,24^. However, the PURE-MRI study revealed poor accuracy of physician urethra contours and very poor inter-physician agreement. Those results call into question both the validity of urethra dose constraints and the feasibility of introducing standardized urethra contouring in general clinical practice. We provide here a multidisciplinary consensus contouring atlas for the urethra (Supplementary Figure 1, Supplementary Figure 2). We also developed an AI tool that outperforms physicians for urethra coverage, covering a median 81% of the reference standard (vs. median 36% across 110 physician contours) without including excessive amounts of non-urethra tissue (measured by Max 2D HD). In a broader multi-center dataset, the AI tool typically covered 89% of the reference standard urethra while straying no more than 2.0 mm from the reference urethra contour.

Dice score (also called Dice similarity coefficient) is a commonly used metric for evaluating segmentation accuracy but can be poorly suited to some applications^25^. The urethra is small in diameter and has a comparatively long course relative to its diameter. It is also poorly visualized in parts of the prostate, and its exact boundary is difficult or impossible to outline even if its location is agreed upon, as noted by our multidisciplinary panel^11^. We therefore proposed percent coverage and Max 2D HD as more appropriate metrics for this case. Coverage of the urethra is of obvious importance if the goal is to avoid overdosing the true urethra during radiation therapy. Conversely, an overly large urethra avoidance structure would unnecessarily limit the dose achievable to some prostate tumors, especially during focal radiation boost^26,27^. Max 2D HD is helpful as an indicator of how overly conservative a urethra contour is in its least accurate slice. Consistent with our reasoning, we found that the AI model was not impressive for Dice score (while still better than physician participants), but the AI tool performed well on percent coverage and Max 2D HD.

Several recent studies have explored the deep learning-based urethra segmentation on MRI or CT^12,13,28^. One such study by Belue et al. reported a model achieving an overall Dice score of 0.61 and typical centerline distance (distance between ground truth and AI segmentation on each slice, averaged across slices of 2.6 mm^13^. It is not valid to compare summary statistics of AI models applied to distinct datasets, but the Dice score in the Belue et al. model is impressive and possibly suggests incorporation of more data for training (Belue et al. had 657 cases for training, compared to the 151 we used), possibly suggesting a larger training dataset would improve our model. Our study, on the other hand, benefitted from including 110 urethra contours made by 62 physicians to directly compare AI performance to human performance on the same patient cases. We also used additional metrics in our study with clinical relevance for radiation treatment planning. Maximum 2D HD is an indicator of error on the worst slice in a patient case and represents the worst-case scenario for designating non-urethra tissue as an avoidance structure that could interfere with focal boost of a nearby tumor. Percent coverage of the urethra is an indicator of how much of the urethra is correctly labeled for sparing (uncovered urethra could lead to urethra doses higher than intended). We are also planning to integrate our urethra model into the clinical workflow by providing physicians with its auto-segmentations, helping them accurately locate the urethra to avoid exceeding dose constraints. This will first be done in two prospective studies, including NCT06990542.

Our study has several limitations. First, establishing a definitive ground truth for urethral contours is inherently challenging. In particular, the upper half of the urethra is often difficult or even impossible to visualize clearly on MRI, making objective delineation unreliable. As a result, the reference standard used in this study—derived from consensus among experienced experts with complementary clinical backgrounds—represents possibly the best feasible approximation at present. Second, we evaluated only a single AI-based segmentation approach. To gain a more comprehensive understanding of the capabilities and limitations of AI tools for urethral segmentation, future studies should include a broader set of algorithms, encompassing both academic methods and commercial solutions. Third, training data came from only one center and 151 patient cases; larger and more diverse training data might improve performance. Fourth, it is unclear how large an error in urethra segmentation must be to have a clinically meaningful impact on patient outcomes, so this cannot be evaluated here. On the other hand, the paucity of clear data on dose-toxicity associations for various urethra-sparing approaches strengthens the rationale for the present study—consistent urethra contours are needed to support robust evidence for better urethra constraints. Our results demonstrate that the AI model developed here already outperforms a large cohort of physicians and offers a promising complementary tool to enhance the accuracy and consistency of urethral contouring in clinical practice.

## Conclusion

We established a multidisciplinary consensus reference-standard dataset for the evaluation of urethra segmentation and provide an atlas for contouring the urethra on MRI. We introduced clinically meaningful metrics, including percent urethra coverage and Max 2D HD, to better reflect segmentation quality in real-world clinical settings. We developed a deep-learning model for urethra segmentation, a challenging and time-consuming task. Our AI tool outperformed most physicians and was comparable to even the best-performing physician participants in the PURE-MRI study. Moreover, its consistent performance across a diverse, multi-center dataset demonstrates overall robustness and generalizability, though there were some errors large enough to require correction. These results suggest a strong potential for AI-based auto segmentation of the urethra to assist, or even augment, physician urethra segmentation during radiation therapy planning, ultimately facilitating investigations of valid dose constraints and helping to reduce the risk of urethral injury.

## Data Availability

All data produced in the present study are available upon reasonable request to the authors

## Notes

### Competing Interest Statement

DJAM reports a clinical advisor role for Stratagen Bio and an ad hoc consultant role for Guerbet and Promaxo. SAW was supported by funding from the ARRS Scholarship for professional development and has received investigator-initiated research grants (paid to the institution) from Siemens. Sophia C. Kamran' spouse is employed by Sanof. AMD is a founder of and holds equity interest in CorTechs Labs and serves on its scientific advisory board. He is also a member of the Scientific Advisory Board of Healthlytix and receives research funding from General Electric Healthcare (GEHC). Rebecca Rakow-Penner Human Longevity Inc: Consultant, Cortechs Labs: Stock options, Curemetrix: Stock options, consultant Imagine Scientific, advisory board. SBIR GE Healthcare, research agreement Bayer consultant. Michael Liss Founder/President of Oncobiomix with no relation to this manuscript. TMS reports honoraria from Varian Medical Systems, WebMD, MJH Life Sciences, GE Healthcare, Blue Earth Diagnostics, and Janssen; he has an equity interest in CorTechs Labs, Inc. and serves on its Scientific Advisory Board; he receives research funding from GE Healthcare and Blue Earth Diagnostics, as well as in-kind research support from Quibim, Inc., both through the University of California San Diego. These companies might potentially benefit from the research results. The terms of this arrangement have been reviewed and approved by the University of California San Diego in accordance with its conflict-of-interest policies. The remaining authors have no conflicts of interest to disclose.

### Funding Statement

National Institutes of Health, Grant/Award Numbers: NIH/NIBIB K08EB026503, NIHUL1TR000100; American Society for Radiation Oncology; Prostate Cancer Foundation; Department of Defense, Grant/Award Number: DOD/CDMRPPC220278

### Author Declarations

The IRBs of University of California San Diego, Massachusetts General Hospital, University of Rochester Medical Center, University of California San Francisco, University of Texas Health Sciences Center San Antonio, and University of Cambridge gave ethical approval for this work.

