## Supplementary Materials for "Urethra contours on MRI: multidisciplinary consensus educational atlas and reference standard for artificial intelligence benchmarking"

| **Characteristics** | **Description** | **Frequency (%), n=62** |
| --- | --- | --- |
| **Country** | Australia | 5 (8%) |
|  | Belgium | 1 (2%) |
|  | Brazil | 1 (2%) |
|  | Cyprus | 2 (3%) |
|  | Denmark | 1 (2%) |
|  | Germany | 5 (8%) |
|  | India | 2 (3%) |
|  | Spain | 1 (2%) |
|  | Switzerland | 1 (2%) |
|  | United Kingdom | 6 (10%) |
|  | United States | 37 (60%) |
| **Setting** | Rural | 1 (2%) |
|  | Suburban | 14 (23%) |
|  | Urban | 47 (76%) |
| **Specialty** | Radiation Oncologist | 43 (69%) |
|  | Radiologist | 12 (19%) |
|  | Urologist | 7 (11%) |
| **GU-Focused Practice** | No | 18 (29%) |
|  | Partially | 25 (40%) |
|  | Completely | 19 (31%) |
| **Clinical Experience** | Still in training | 14 (23%) |
|  | <5 years | 12 (19%) |
|  | 5-10 years | 11 (18%) |
|  | > 10 years | 25 (40%) |

Supplementary Table 1. Summary characteristics of physician participants in the PURE-MRI study.

| **Scanner** | **Dataset 1** |
| --- | --- |
| **Pulse sequence** | Fast Spin Echo (FSE) |
| **TR (ms)** | 3276-3615 |
| **TE (ms)** | 98-172 |
| **FOV (mm)** | 160 x 160, 180 x 180 |
| **Matrix [resampled dimensions]** | 360 x 224 [400 x 256] |
| **Slices** | 32 |
| **Slice Thickness (mm）** | 3 |
| **Field Strength (T)** | 3 |

Supplementary Table 2. MRI acquisition parameters *T_2_*-weighted sequences for Dataset 1. TR: repetition time. TE: echo time. FOV: field-of-view. FSE: fast spin echo.

| **Scanner** | **Imaging Center 1*** | **Imaging Center 2** | **Imaging Center 3** | **Imaging Center 4** | **Imaging Center 5** | **Imaging Center 6** |
| --- | --- | --- | --- | --- | --- | --- |
| **Pulse sequence** | Fast Spin Echo (FSE) | Turbo Spin Echo (FSE) | Fast Spin Echo (FSE) | Fast Spin Echo (FSE) | Fast Spin Echo (FSE) | Turbo Spin Echo (FSE) |
| **TR (ms)** | 5300 | 4800 | 4800 | 2964 | 3130 | 4710 |
| **TE (ms)** | 100 | 104 | 104 | 150 | 98 | 100 |
| **FOV (mm)** | 200 x 200 | 180 x 180 | 180 x 180 | 220 x 220 | 180 x 180 | 180 x 180 |
| **Matrix [resampled dimensions]** | 320 x 320 [512 x 512] | 384 x 365 [384 x 384] | 384 x 365 [384 x 384] | 512 x 512 [320 x 320] | 448 x 256 [512 x 512] | 240 x 320 [320 x 320] |
| **Slices** | 32 | 32 | 32 | 36 | 30 | 30 |
| **Slice Thickness (mm）** | 3 | 3 | 3 | 3 | 3 | 3 |
| **Field Strength (T)** | 3 | 3 | 3 | 3 | 3 | 3 |

Supplementary Table 3. MRI acquisition parameters *T_2_*-weighted sequences for each cohort of Dataset 2. TR: repetition time. TE: echo time. FOV: field-of-view. FSE: fast spin echo. *One of the ten patients from Imaging Center 1 was scanned with a TR of 7000 ms and FOV 240 x 240 mm.

| **Profession** | **Dice** | | | **Coverage (%)** | | | **HD (mm)** | | |
| --- | --- | --- | --- | --- | --- | --- | --- | --- | --- |
| **Profession** | median | | | median | | | median | | |
|  | min | IQR | max | min | IQR | max | min | IQR | max |
| **All Experts (n=62)** | 0.33 | | | 36 | | | 1.6 | | |
|  | 0.03 | 0.23-0.42 | 0.69 | 3 | 25-59 | 96 | 1.0 | 1.2-1.7 | 2.3 |
| **Radiologists (n=12)** | 0.24 | | | 33 | | | 1.7 | | |
|  | 0.06 | 0.22-0.35 | 0.56 | 3 | 25-41 | 75 | 1.1 | 1.5-2.0 | 2.2 |
| **Radiation Oncologists (n=43)** | 0.36 | | | 36 | | | 1.5 | | |
|  | 0.03 | 0.26-0.47 | 0.69 | 3 | 25-61 | 91 | 1.0 | 1.1-1.7 | 2.2 |
| **Urologists (n=7)** | 0.27 | | | 56 | | | 1.7 | | |
|  | 0.10 | 0.21-0.30 | 0.35 | 6 | 34-80 | 96 | 1.1 | 1.6-2.1 | 2.3 |

Supplementary Table 4. Performance of human expert participants with different profession on urethra segmentation on Dataset 1.

| 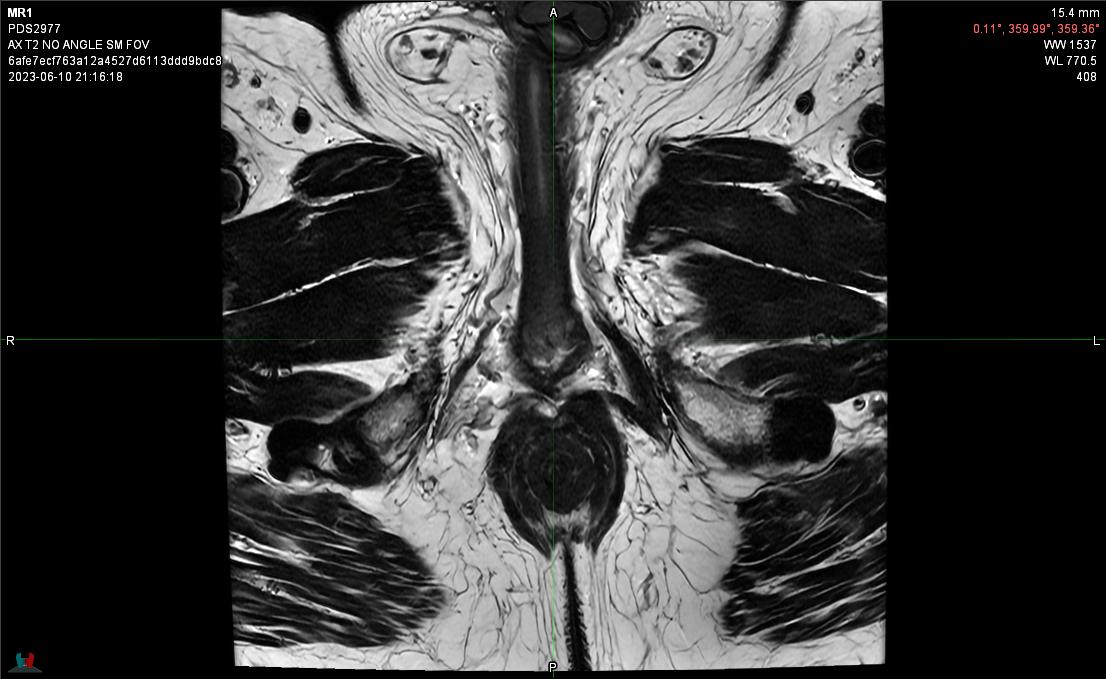  Axial View Slice 1 | 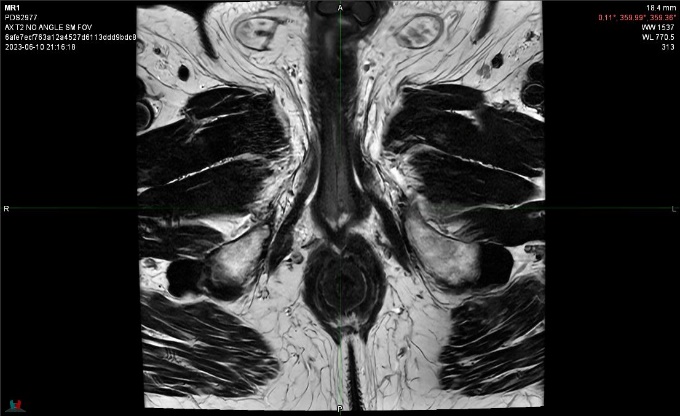  Axial View Slice 2 |
| --- | --- |
| 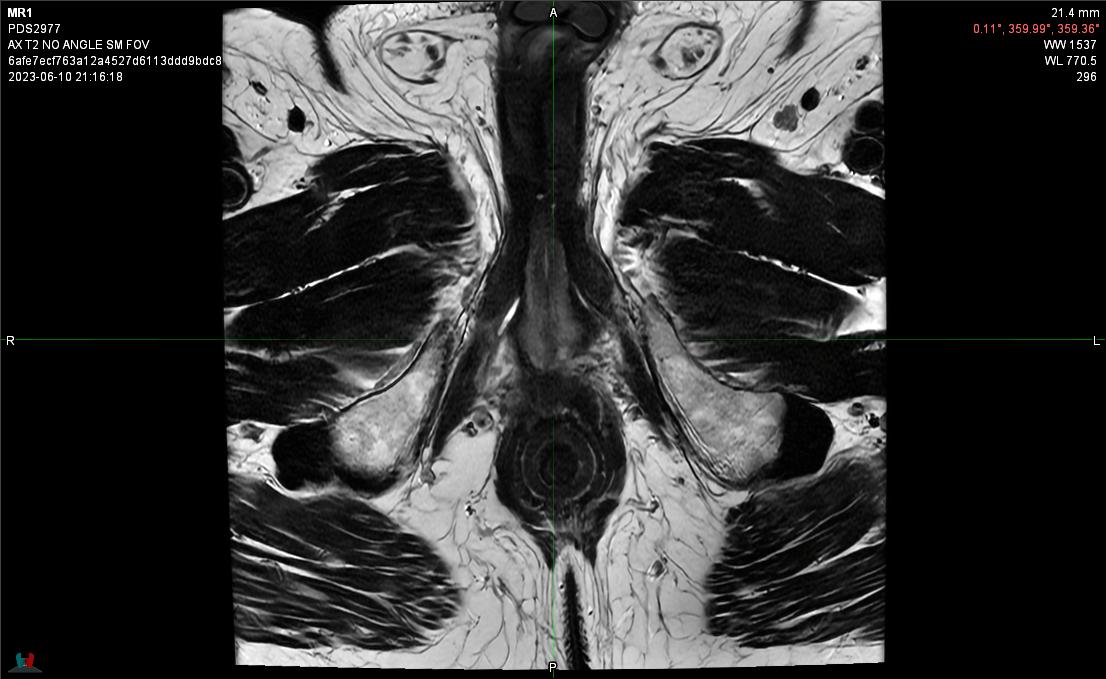  Axial View Slice 3 | 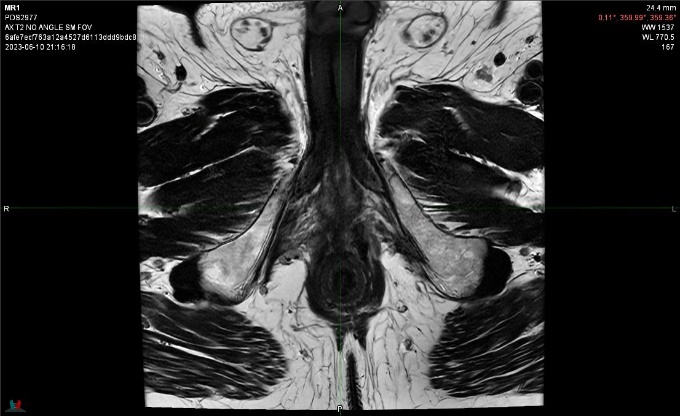  Axial View Slice 4 |
| 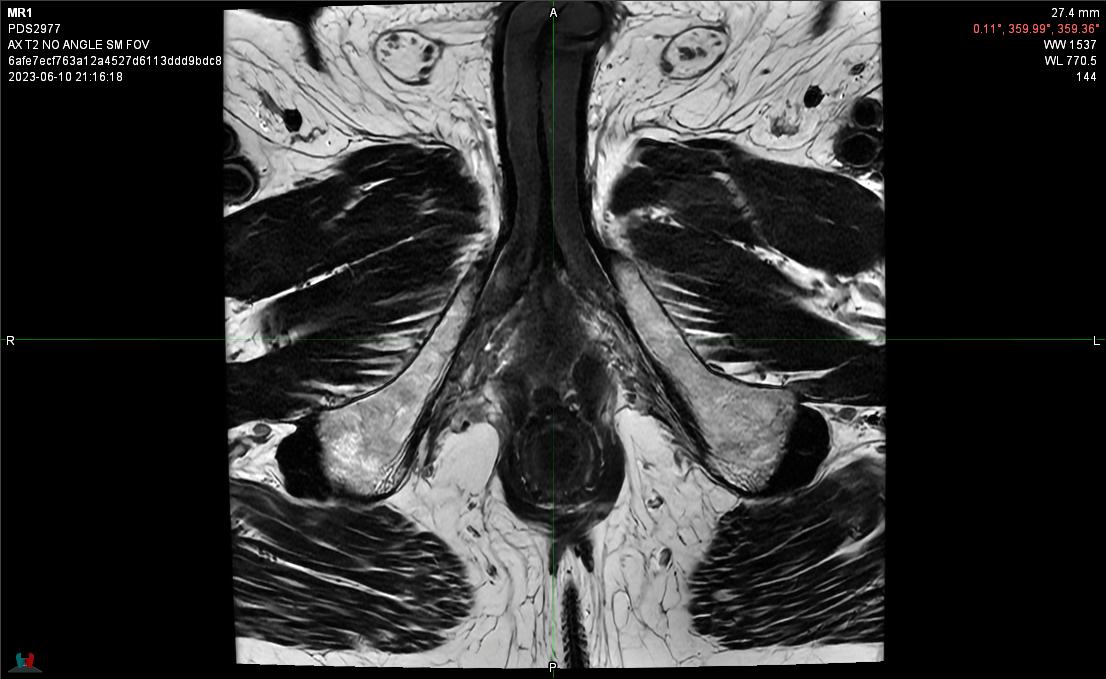  Axial View Slice 5 | 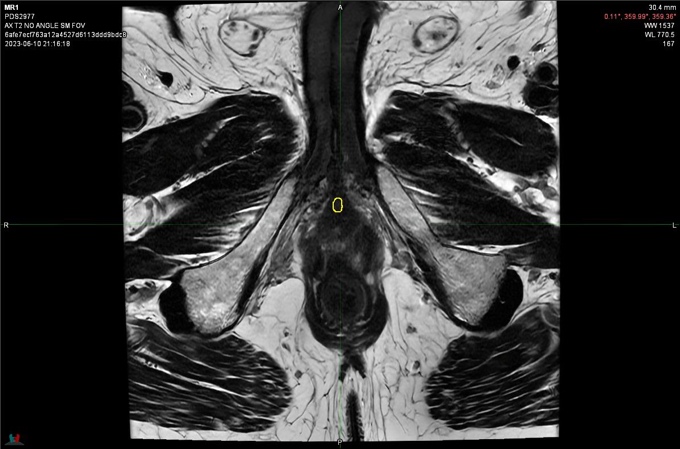  Axial View Slice 6 |
| 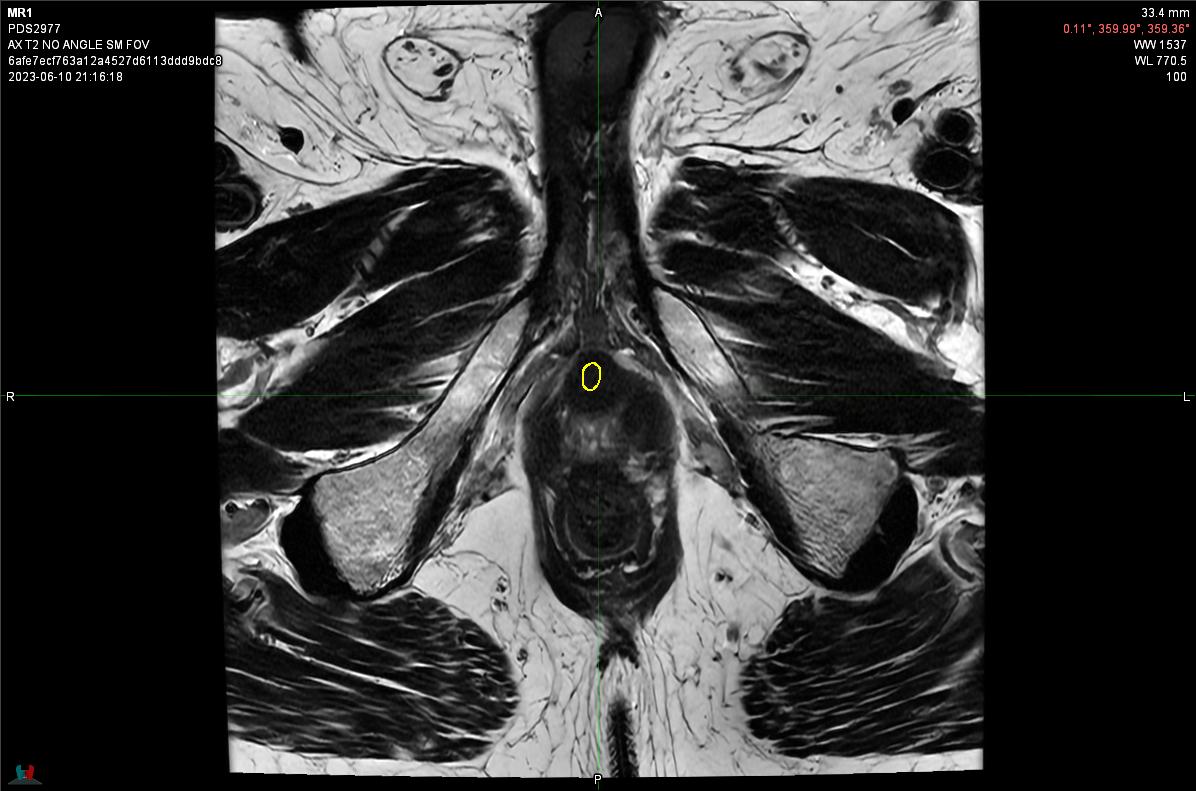  Axial View Slice 7 | 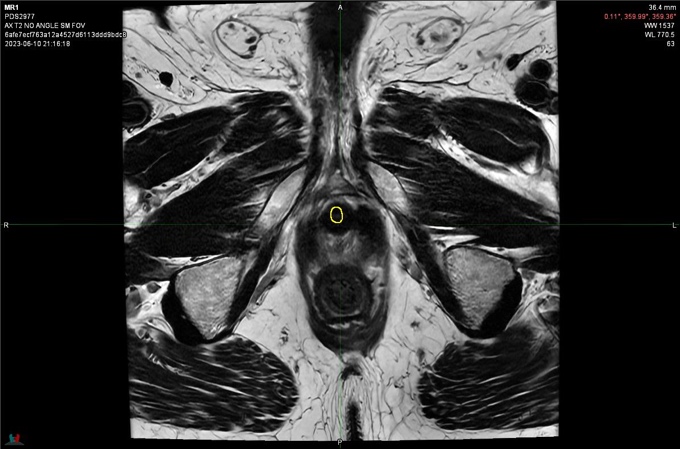  Axial View Slice 8 |
| 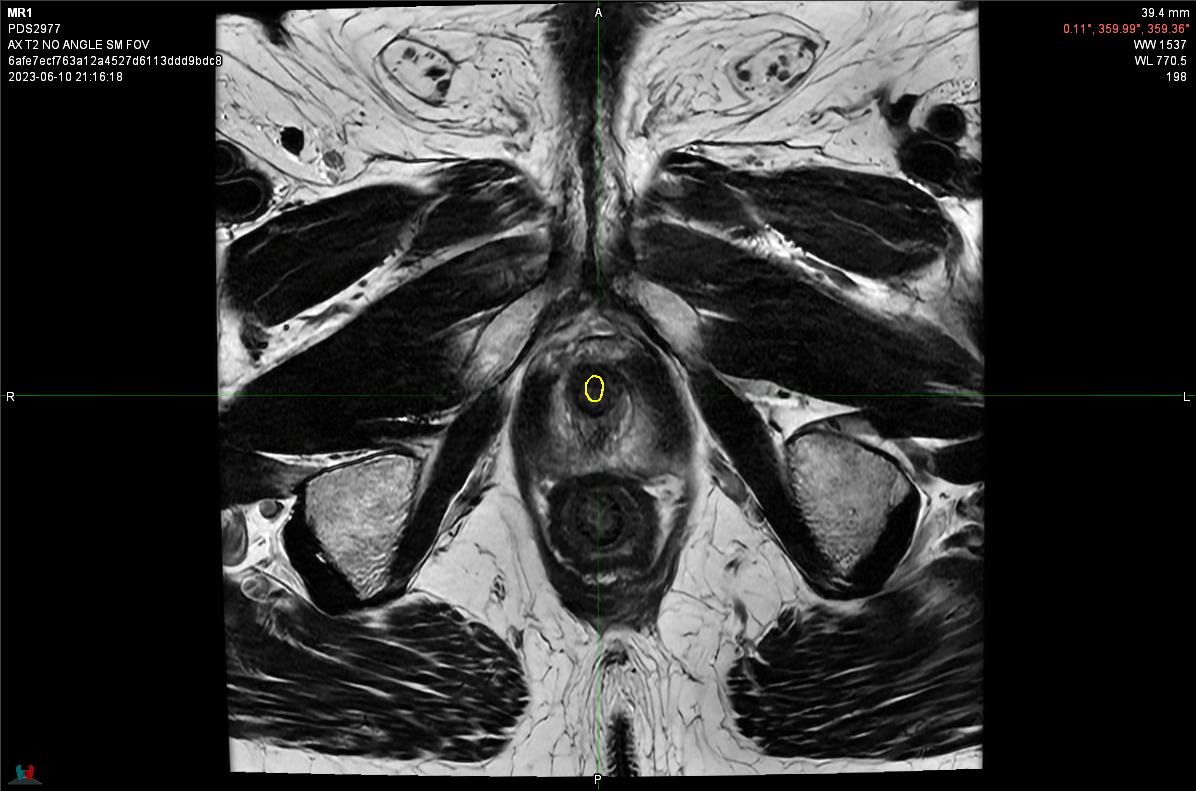  Axial View Slice 9 | 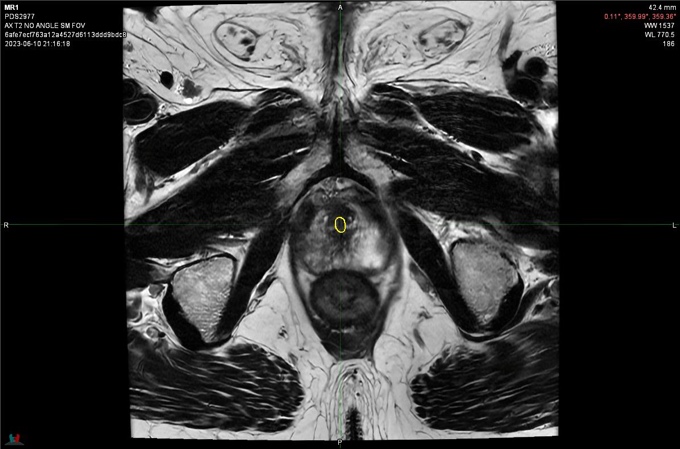  Axial View Slice 10 |
| 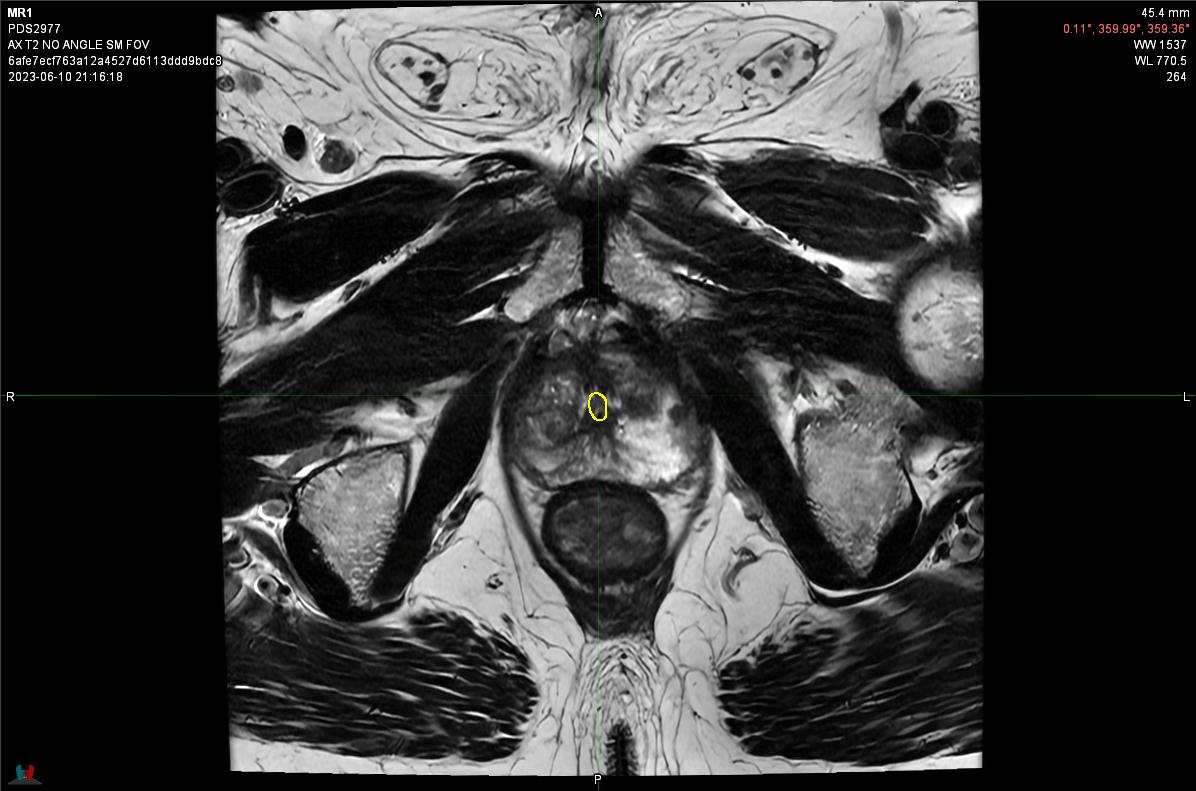  Axial View Slice 11 | 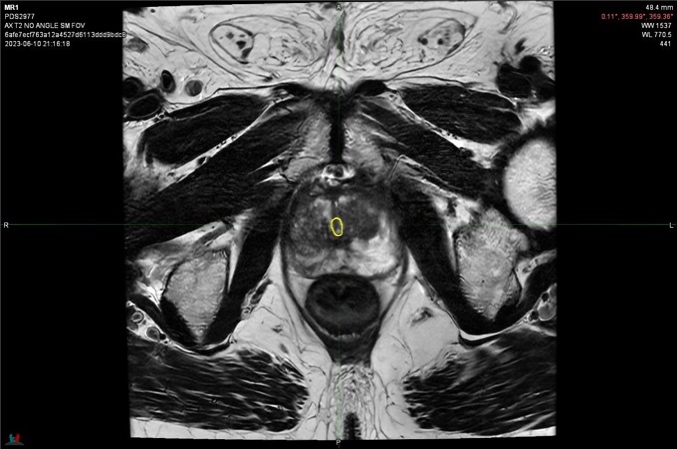  Axial View Slice 12 |
| 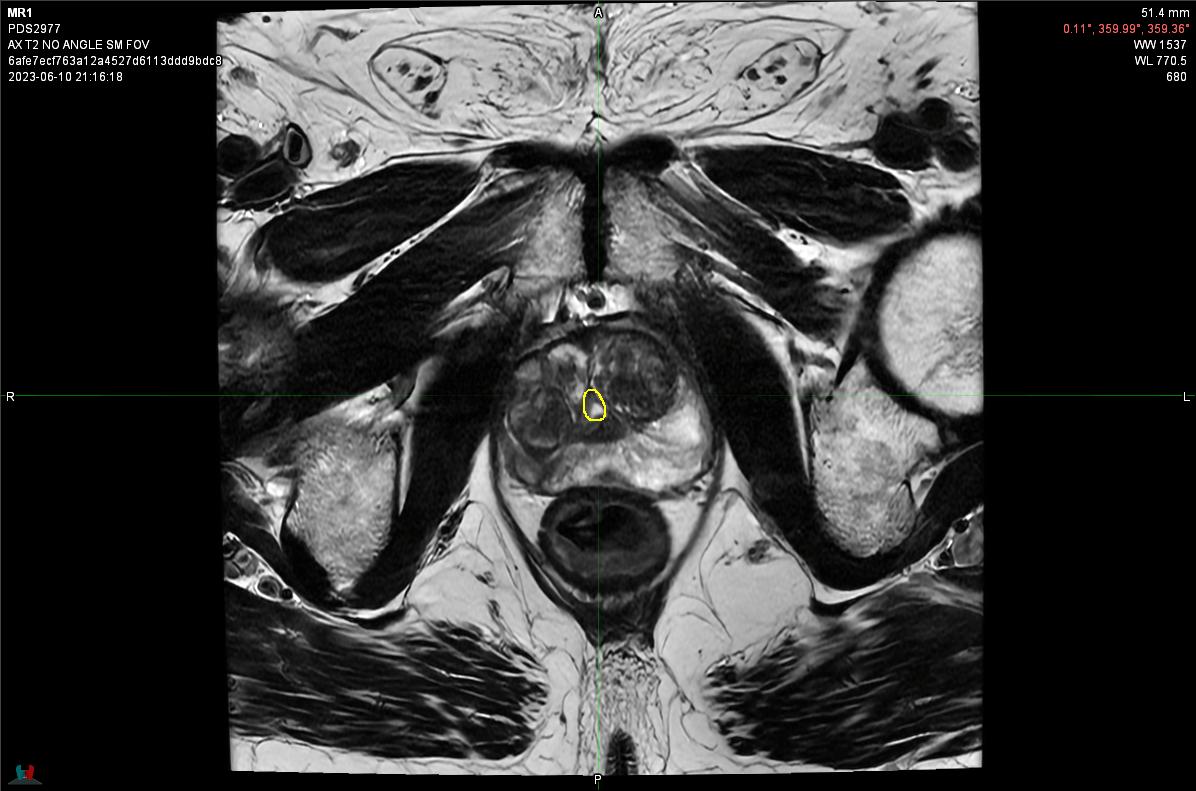  Axial View Slice 13 | 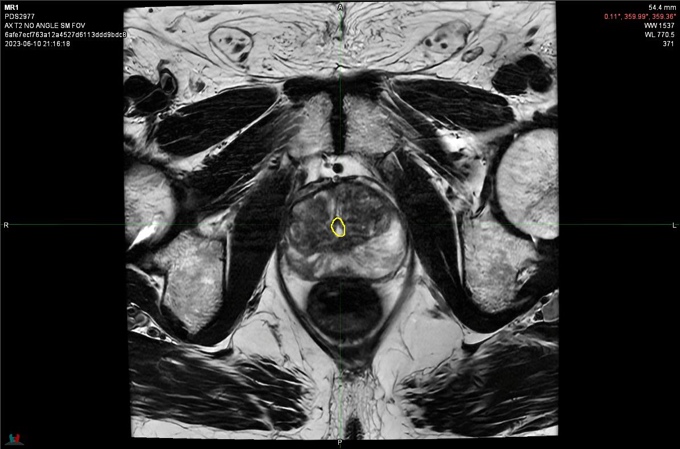  Axial View Slice 14 |
| 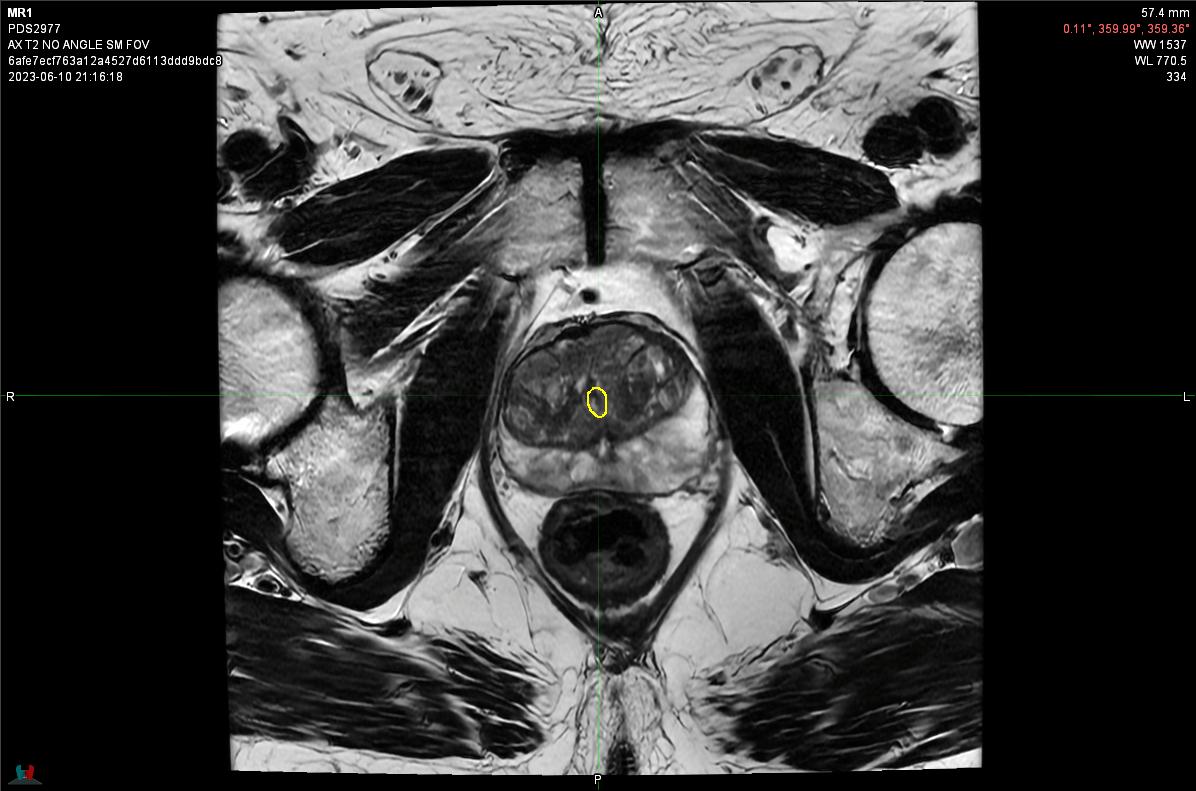  Axial View Slice 15 | 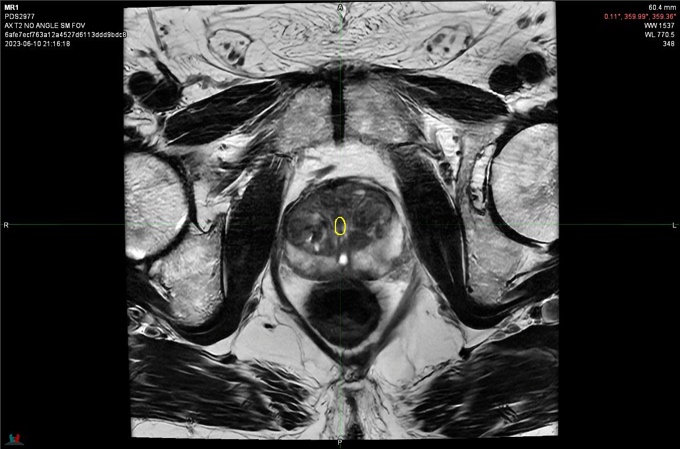  Axial View Slice 16 |
| 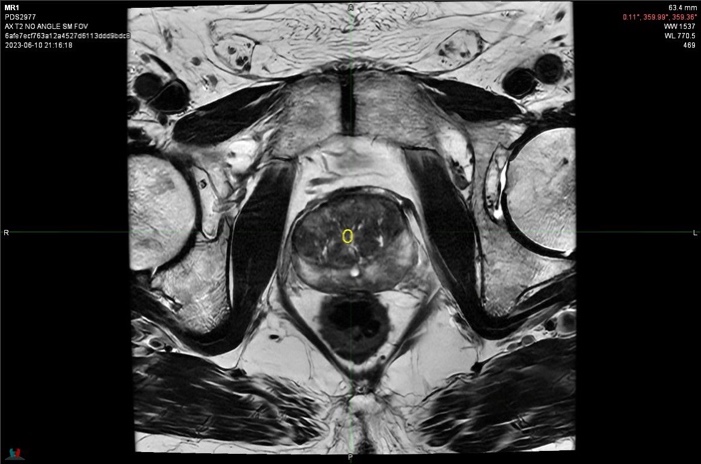  Axial View Slice 17 | 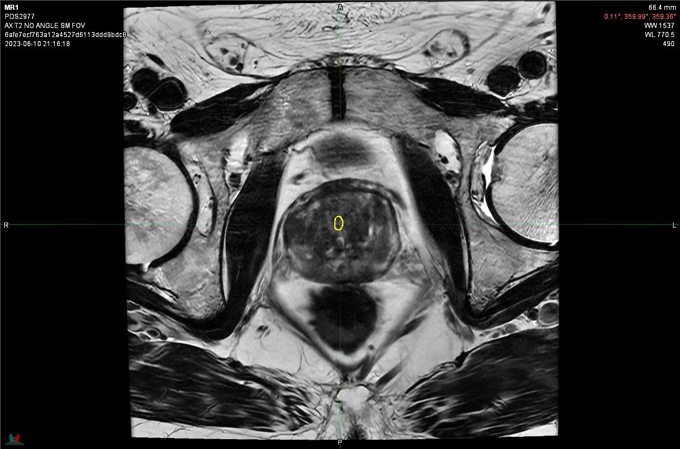  Axial View Slice 18 |
| 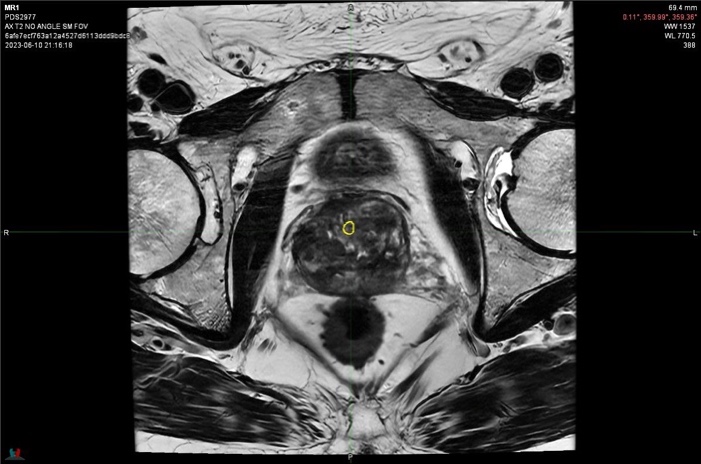  Axial View Slice 19 | 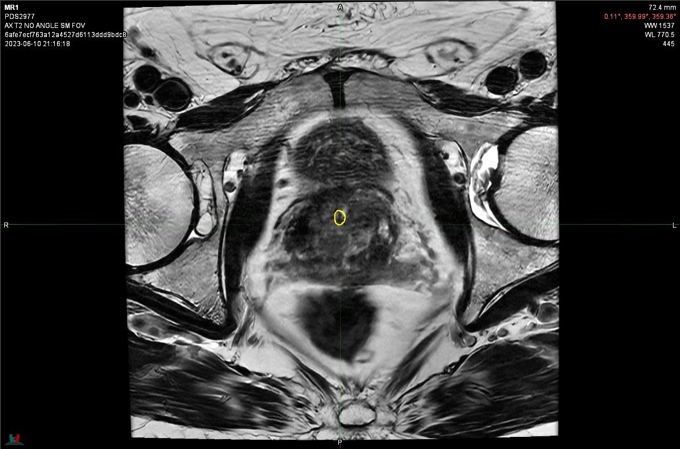  Axial View Slice 20 |
| 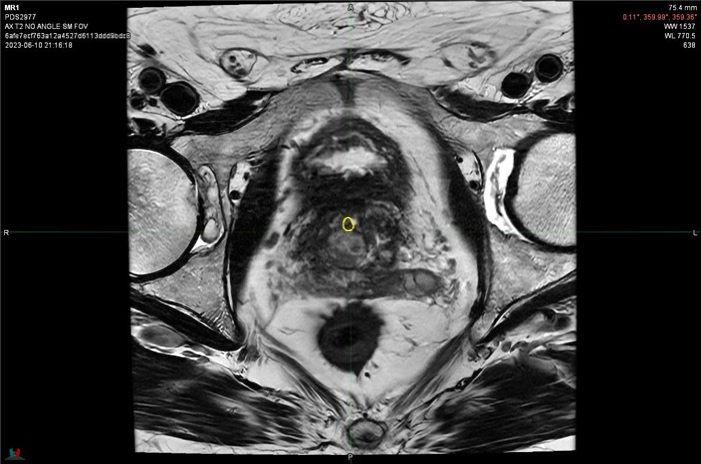  Axial View Slice 21 | 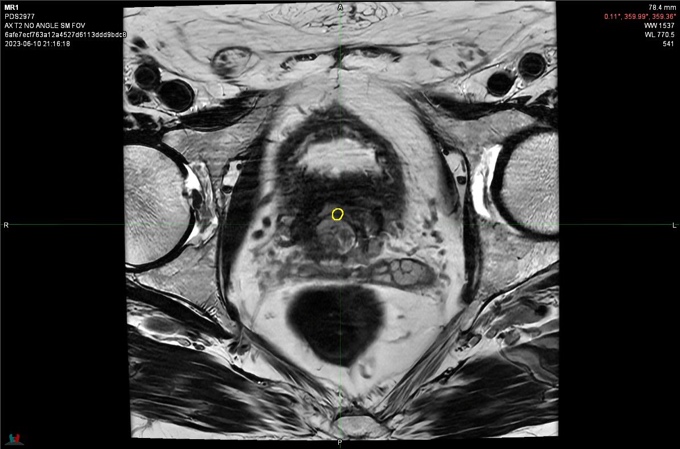  Axial View Slice 22 |
| 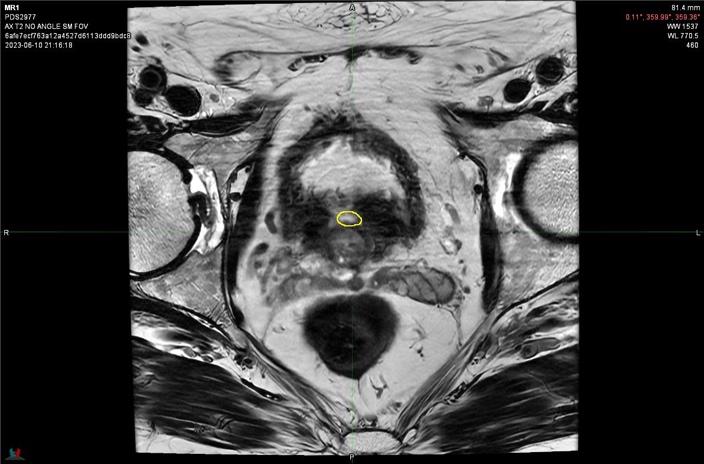  Axial View Slice 23 | 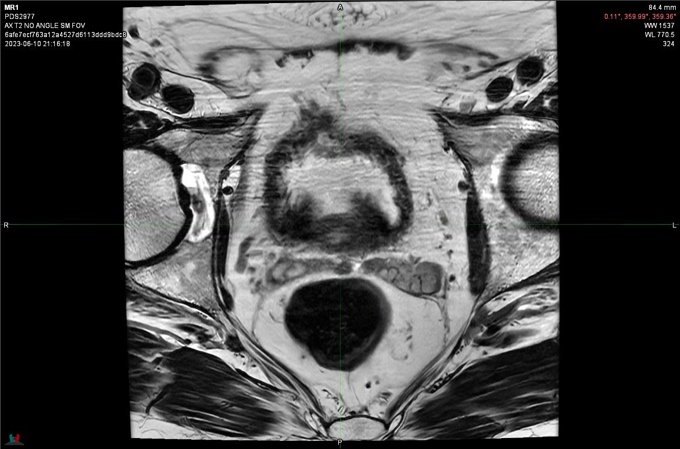  Axial View Slice 24 |
| 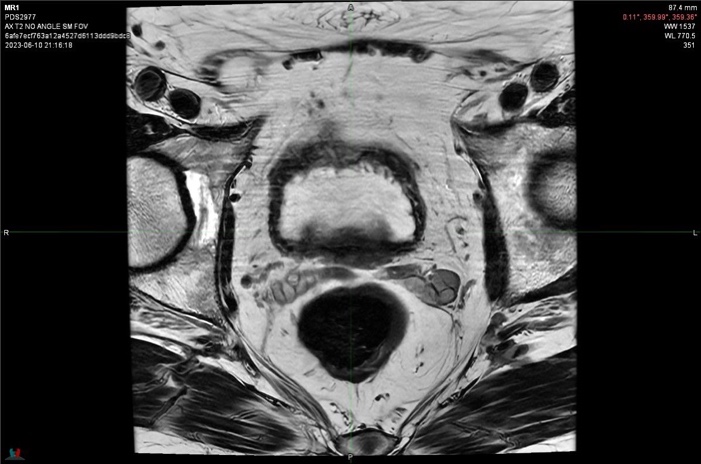  Axial View Slice 25 | 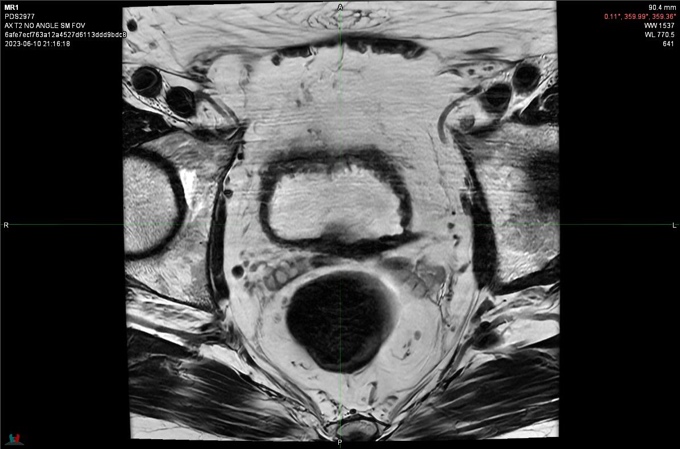  Axial View Slice 26 |
| 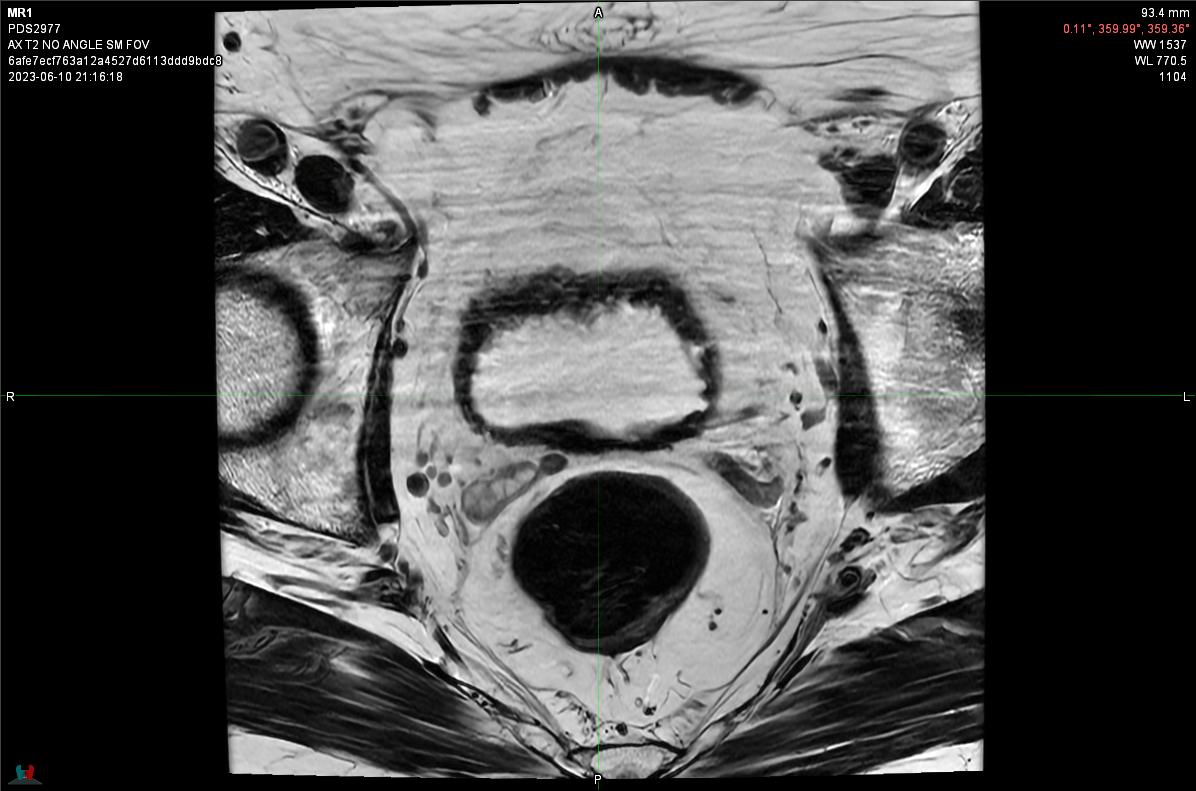  Axial View Slice 27 | 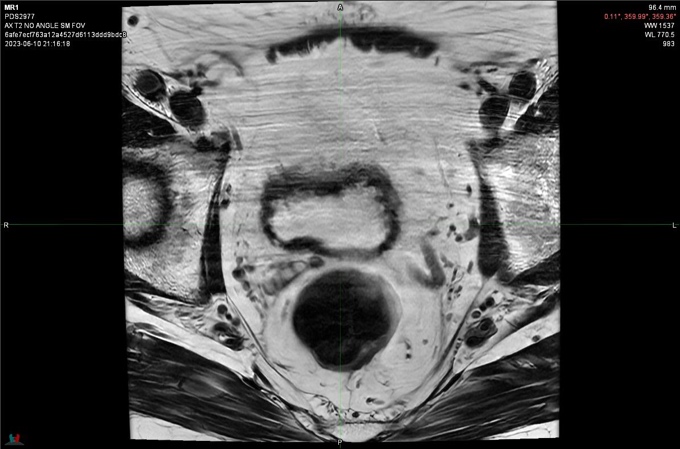  Axial View Slice 28 |
| 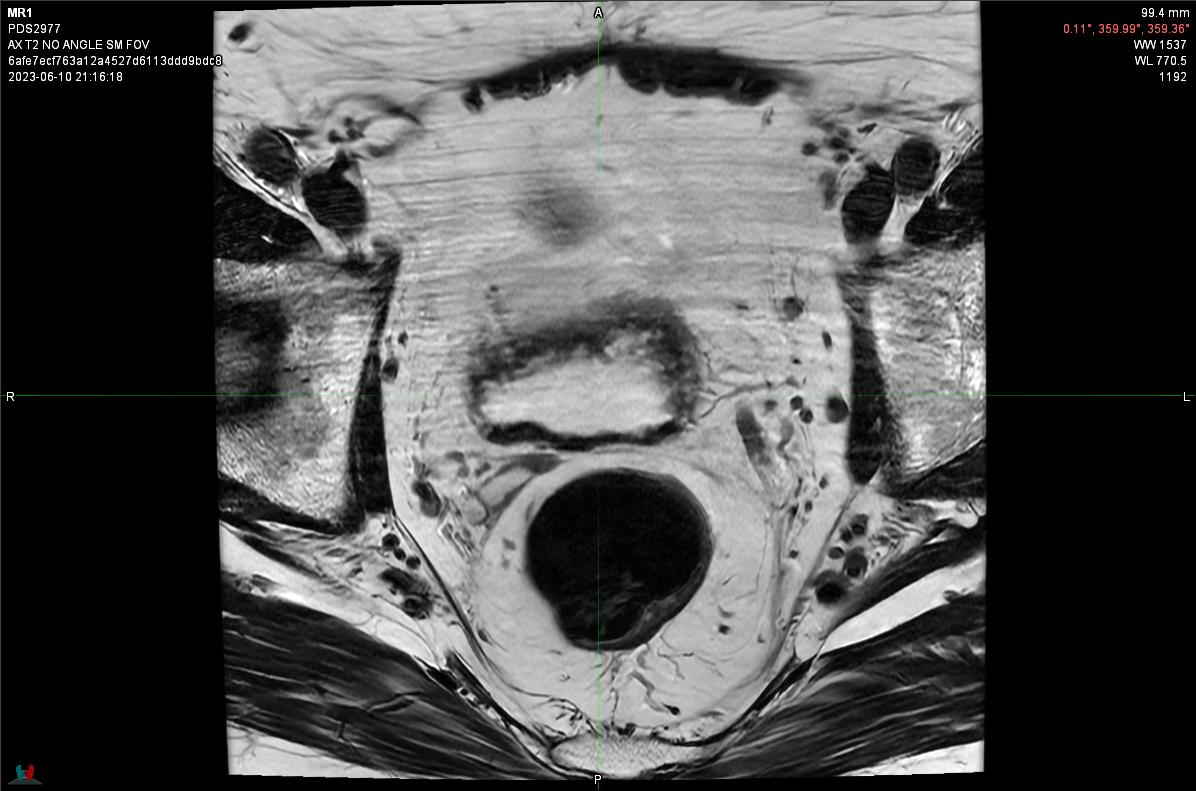  Axial View Slice 29 | 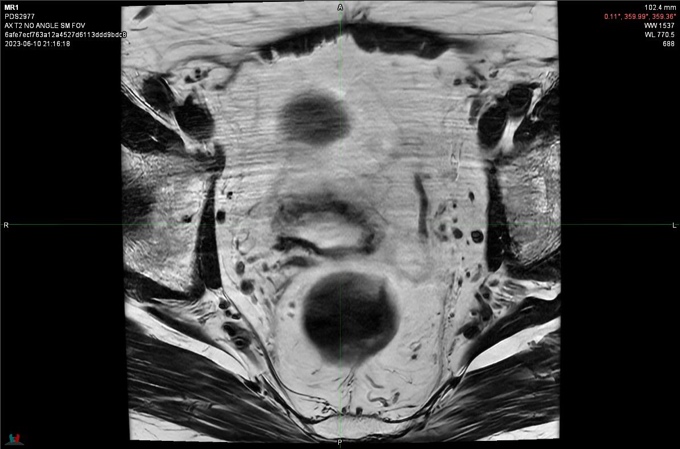  Axial View Slice 30 |
| 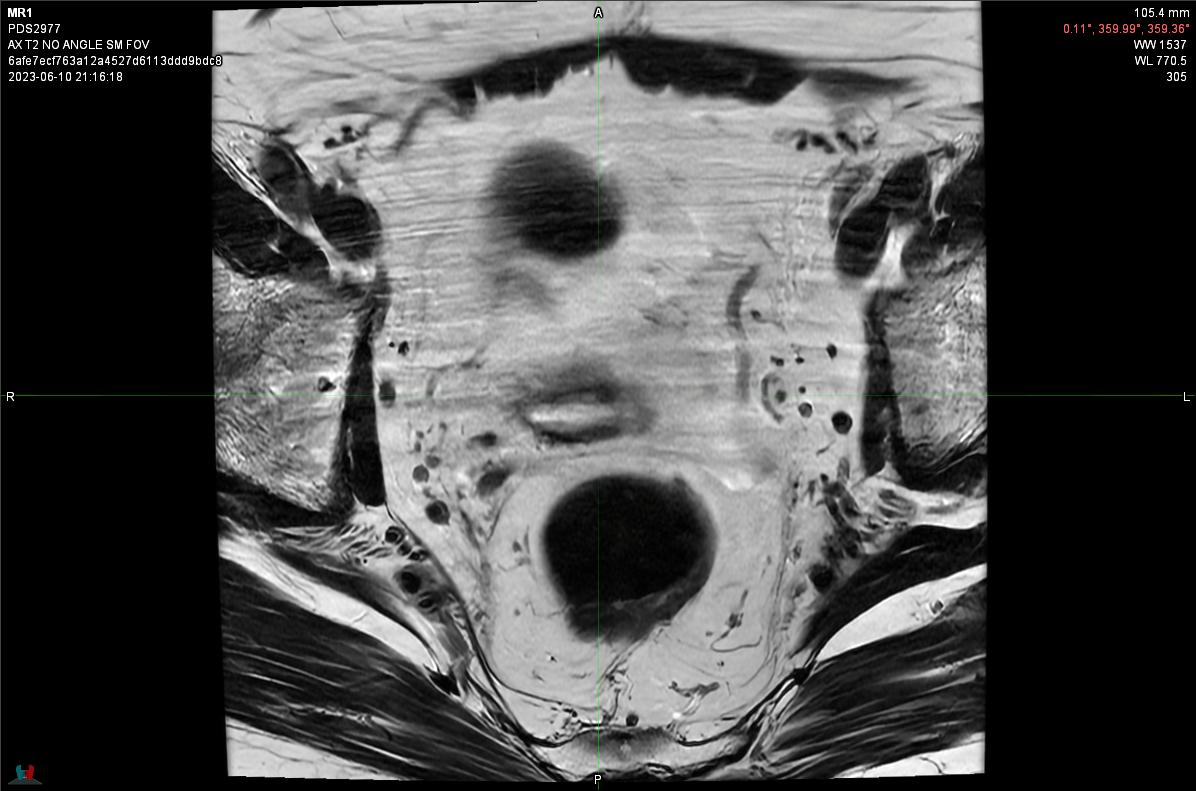  Axial View Slice 31 | 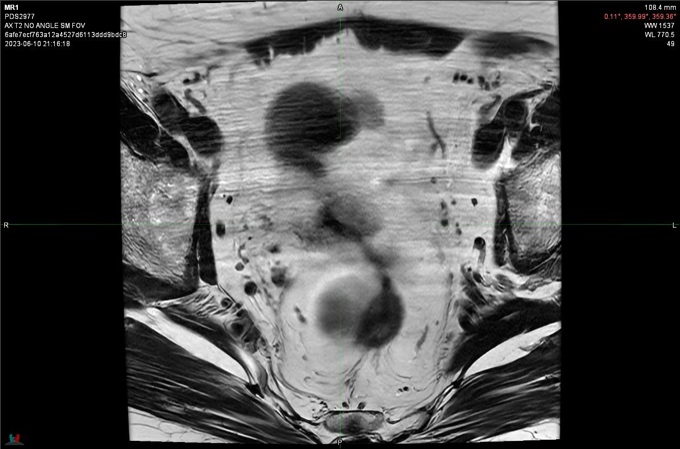  Axial View Slice 32 |
| 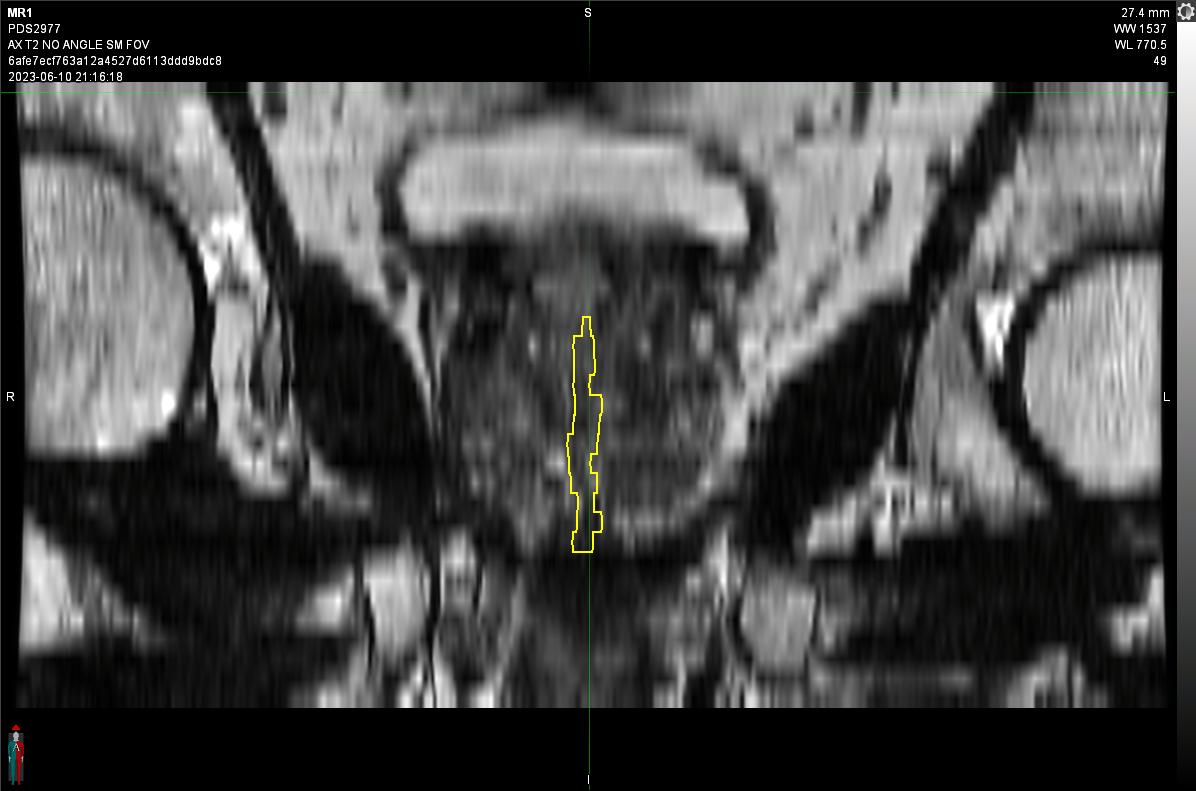Mid Coronal View | 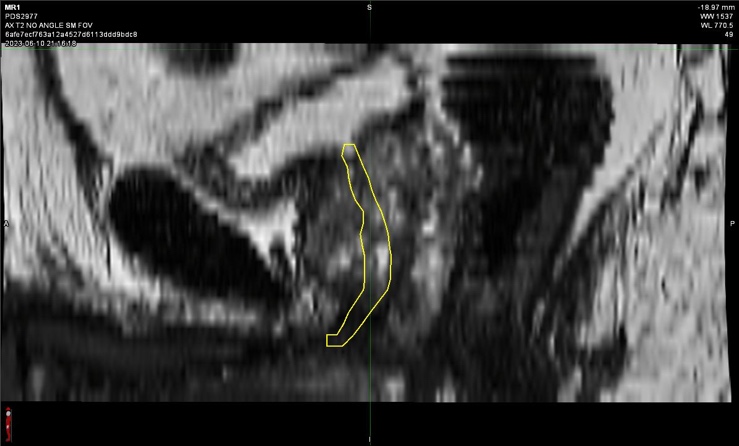Mid Sagittal View |

Supplementary Figure 1. Urethra contouring guide/atlas. A multidisciplinary panel generated this consensus urethra contour on MRI for a representative patient case. The expert contour is shown as yellow contour on axial *T_2_*-weighted slices.

| Axial *T_2_*-weighted Slices | Annotations for Contouring |
| --- | --- |
| 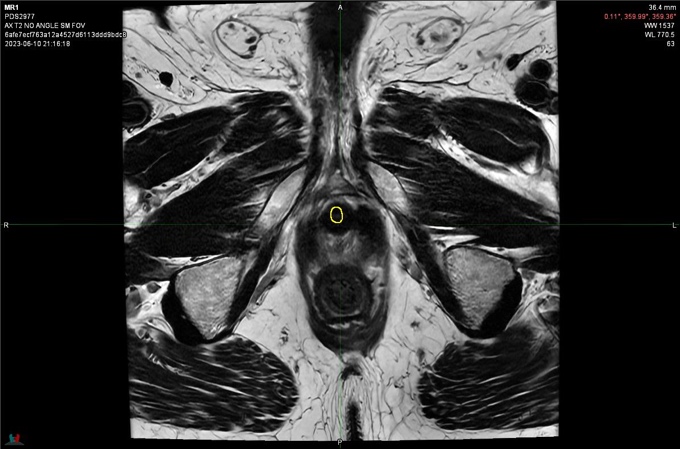  Axial View Slice 8 | Membranous urethra extends from prostatic urethra to inferior aspect of urogenital diaphragm, ending just superior to the penile bulb. |
| 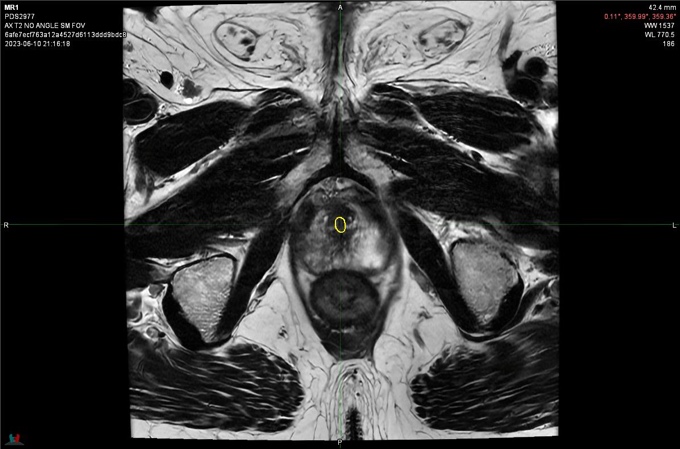  Axial View Slice 10 | Inferior prostatic urethra: continuous with membranous urethra and often moderately hyperintense, just anterior to an inverted “V” of hyperintensity that can extend superiorly to the verumontanum. |
| 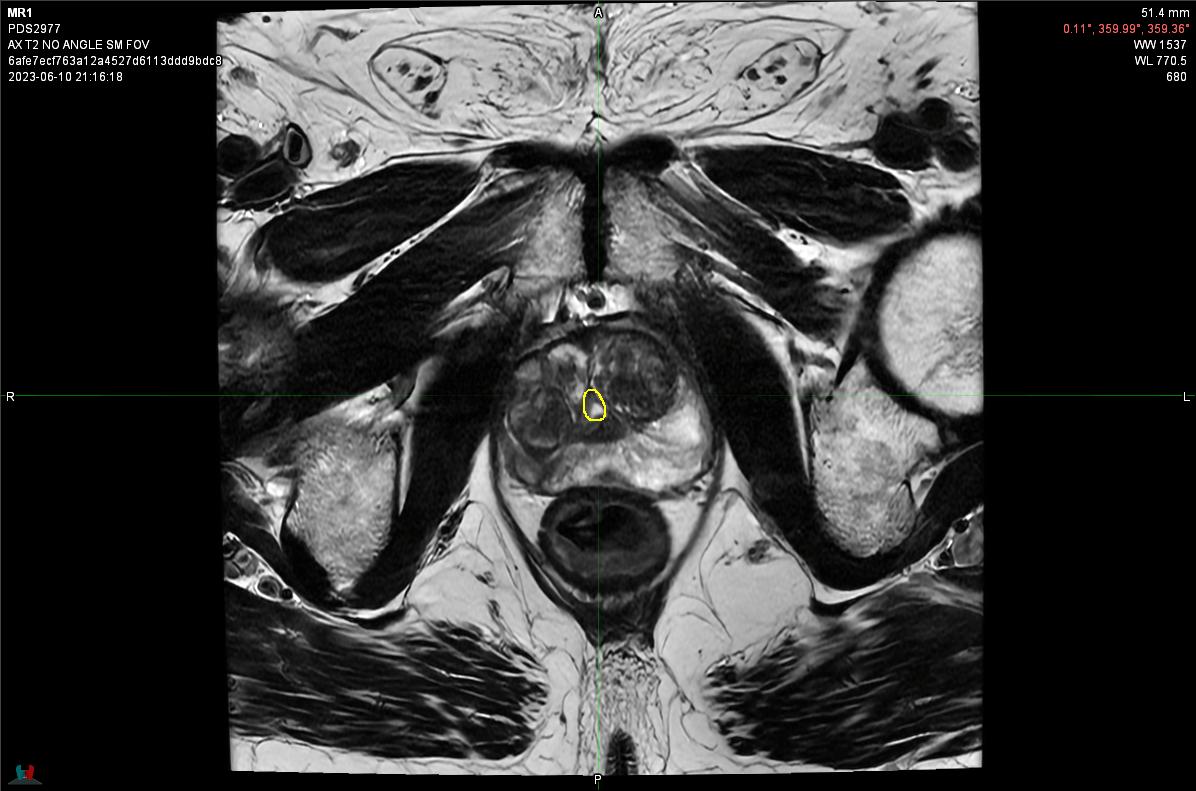  Axial View Slice 13 | Example of urethra at level of verumontanum. Superior to this, the urethra often becomes difficult to visualize. |
| 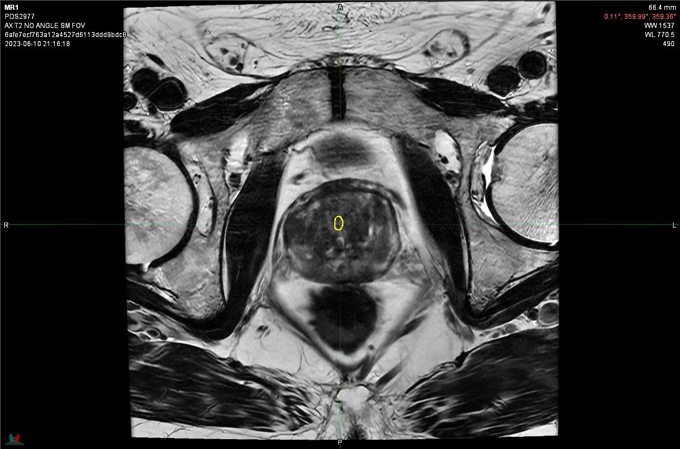  Axial View Slice 18 | Midgland prostatic urethra: The urethra can be difficult to visualize in the upper midgland. It can be helpful to start at apex and track as far superiorly as possible then skip to the prostate base to identify the bladder neck and track the urethra as far inferiorly as possible before returning to the midgland. Consider viewing all three planes while delineating the urethra in the midgland. The urethra may not remain in the mid-sagittal plane because of benign prostatic hyperplasia that causes asymmetry of the prostate. |
| 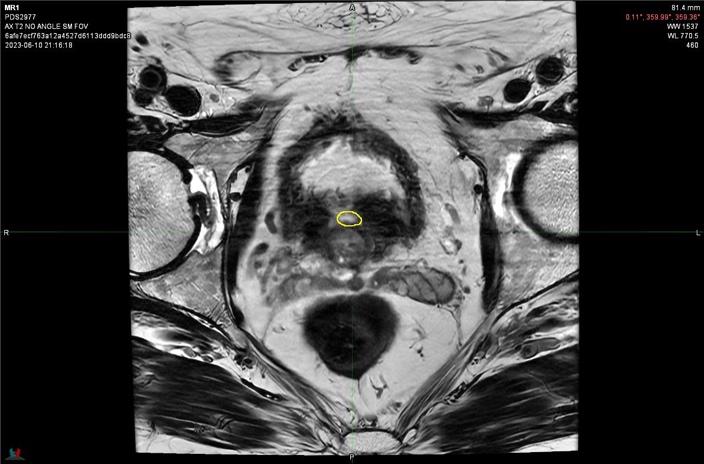  Axial View Slice 23 | Superior prostatic urethra: identify region of intense hyperintensity (urine) where the bladder neck inserts into the prostate base and trace inferiorly. The superior origin of the urethra must be continuous with the bladder neck; it is often not at the superior tip of the prostate but is on the anterior aspect of the gland. |

Supplementary Figure 2. Annotations for urethra contouring. The expert contour is shown as yellow contour on axial *T_2_*-weighted slices on the right column of this figure and annotations for urethra contouring on the left column.

|  | Axial | Coronal | Sagittal |
| --- | --- | --- | --- |
| **A** | 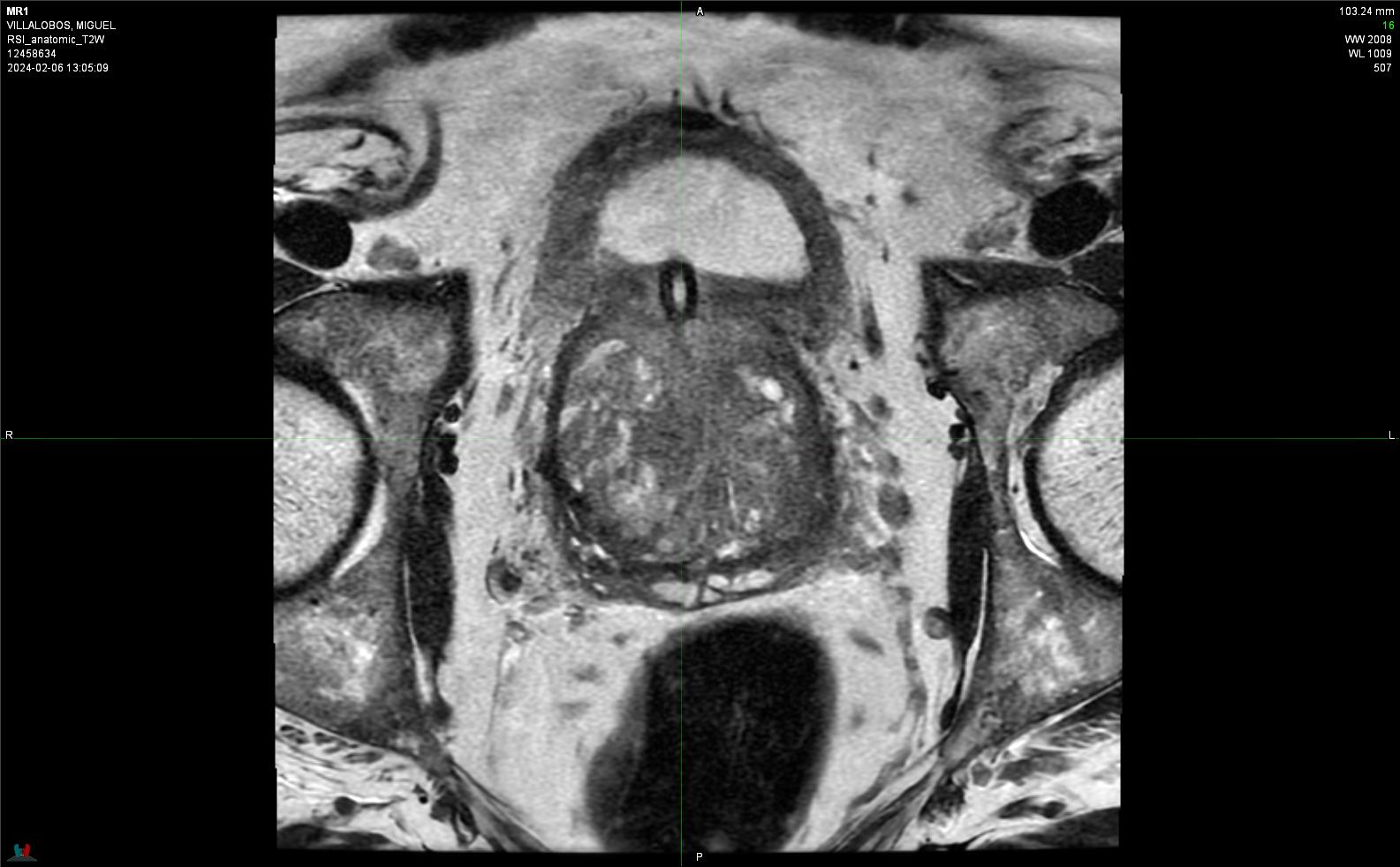 | 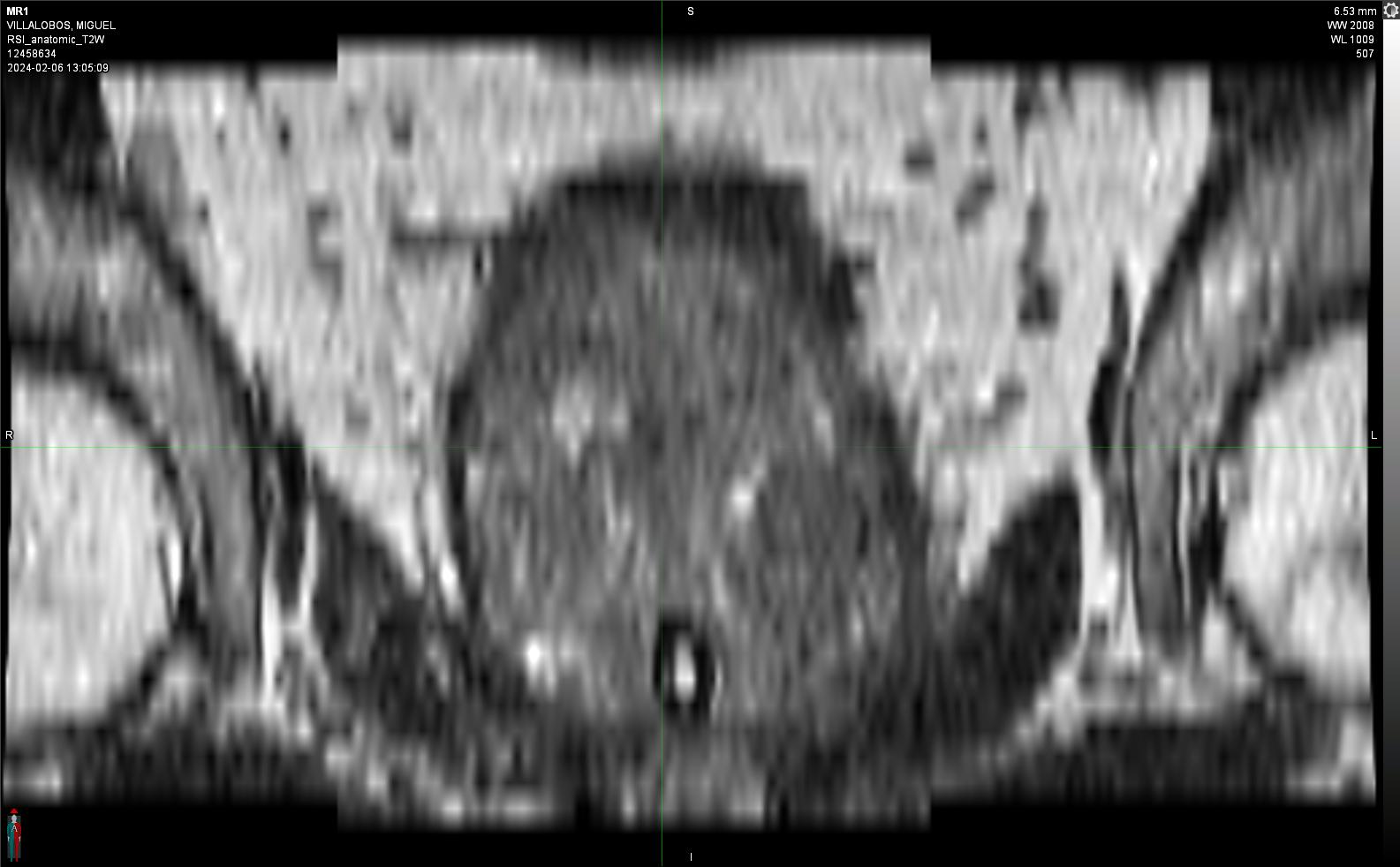 | 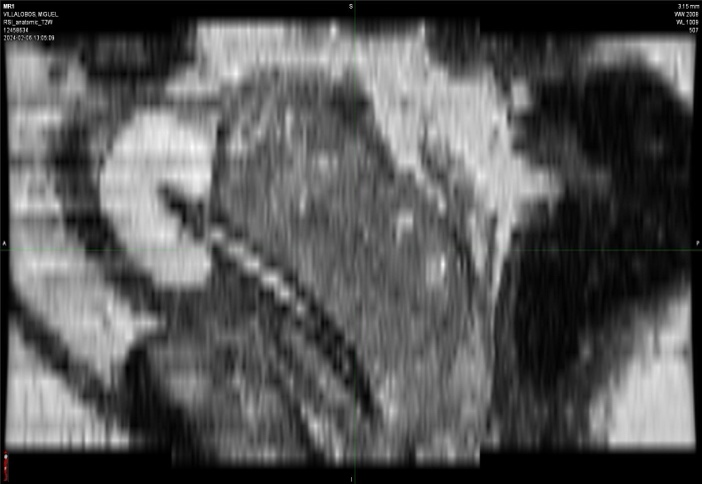 |
| **B** | 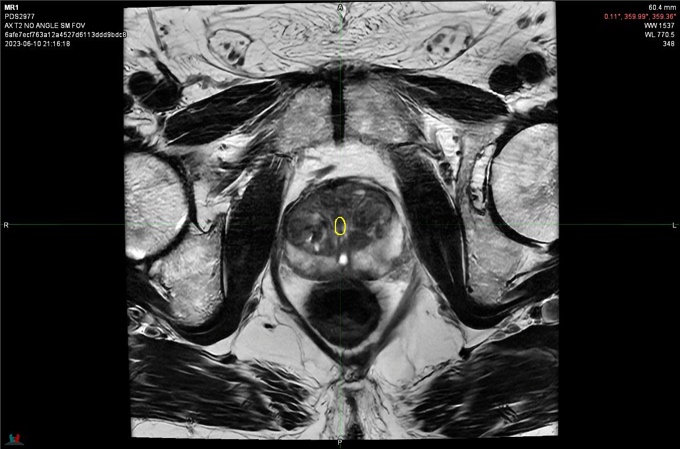 | 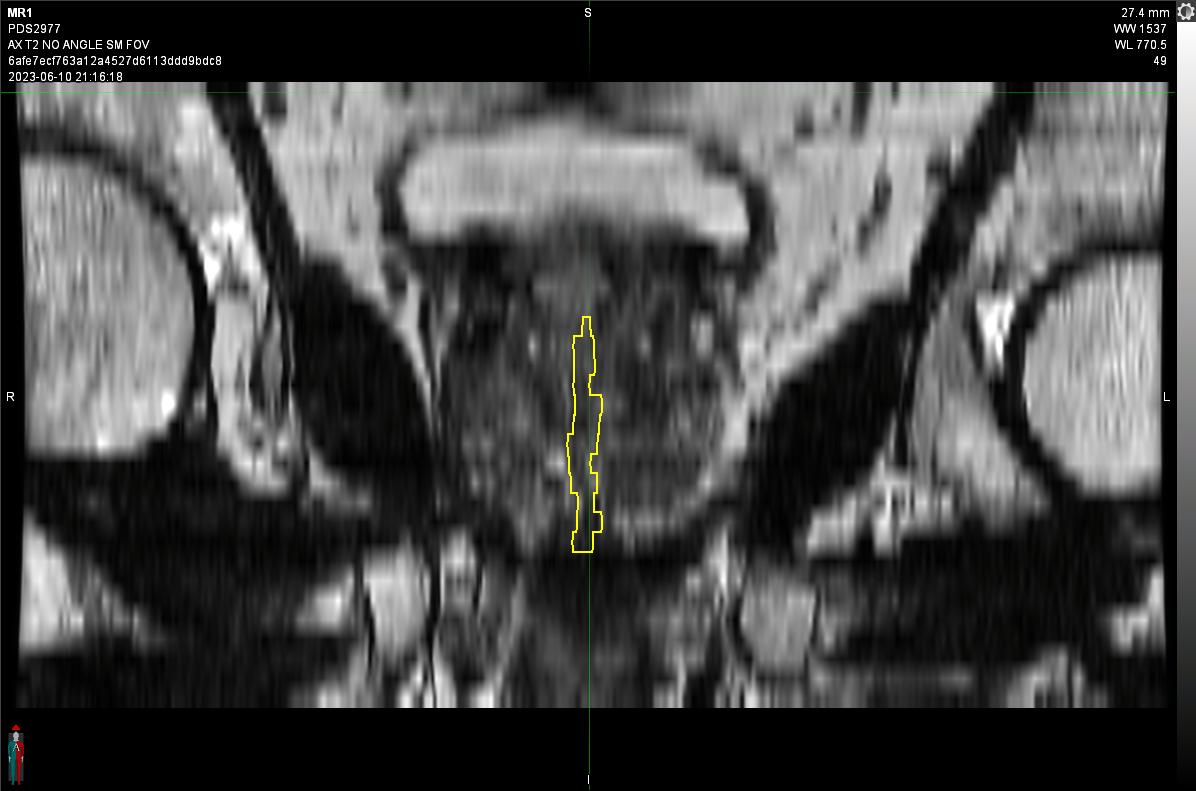 | 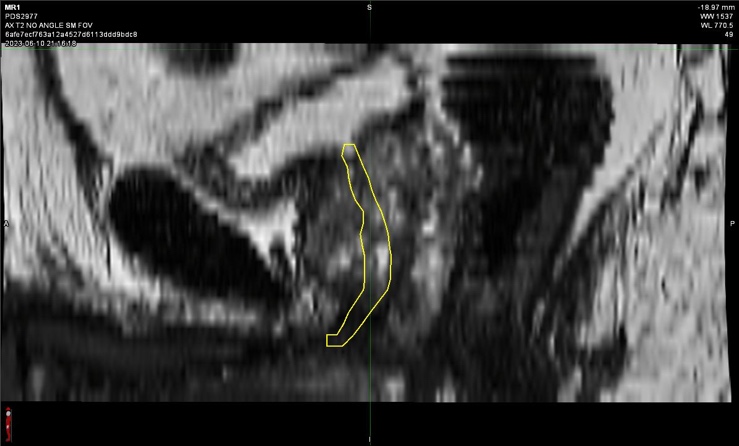 |

Supplementary Figure 3. Example of case with and without catheter. **3A** is an example of a patient with a catheter, the catheter is shown in the red box. **3B** is an example of a patient without a catheter, the expert-defined urethra contour is shown as yellow contour.
